# Whole-genome sequencing enables improved AML diagnostics and refines assessment of *TP53* allelic state and complex karyotype

**DOI:** 10.64898/2026.11.13.26368028

**Authors:** Petr Stanek, Christina Orsmark-Pietras, Rasmus Henningsson, Marianne Rissler, Niklas Landberg, Helena Ågerstam, Anna Staffas, Linda Fogelstrand, Lars Palmqvist, Gisela Barbany, Mattis Knulst, Gunnar Juliusson, Vladimir Lazarevic, Valtteri Wirta, Lucia Cavelier, Richard Rosenquist, Henrik Lilljebjörn, Thoas Fioretos

## Abstract

Accurate genetic characterization is central to modern acute myeloid leukemia (AML) diagnostics, yet current standard-of-care (SoC) testing relies on multiple complementary assays. We compared paired tumor–normal whole-genome sequencing (WGS) with SoC diagnostics in a population-based cohort of 109 adult AML patients, assessing variant detection, disease classification according to ICC 2022 and WHO 2022, and risk stratification according to ELN 2022. WGS showed high concordance with SoC across variant classes. When considering all reported SoC variants together with high-confidence WGS variants as true positives, WGS showed equal to higher sensitivity than SoC for single-nucleotide variants and small insertions/deletions (97% vs 92%), structural variants (SVs; 100% vs 87%), and copy-number alterations (CNAs; 88.5% both). WGS identified clinically important lesions missed by SoC in 16 cases (15%), including *FLT3*-ITDs and *KMT2A*, *NUP98* and *MECOM* rearrangements. Classification and risk assignments were 95–96% concordant, with WGS-driven reclassifications in 4-5% of cases, primarily through detection of cryptic SVs and myelodysplasia-related lesions. Integrated analysis of genomic alterations, including copy-neutral loss-of-heterozygosity, refined *TP53* allelic state assessment and demonstrated increased genomic complexity in AML with biallelic *TP53* alterations. We developed a supervised machine-learning model for complex karyotype (CK) probability, based on WGS-derived copy-number and SV burden, enabling automated identification of CK cases from WGS data. Collectively, these results support paired tumor– normal WGS as a single comprehensive assay well suited for AML diagnostics according to current classification and risk-stratification systems, while refining assessment of *TP53* allelic state and karyotypic complexity.

## INTRODUCTION

Acute myeloid leukemia (AML) is a genetically heterogeneous malignancy characterized by diverse genomic alterations that critically impact diagnosis, prognosis, and treatment selection.^1–6^ Given its complexity, accurate genetic classification and risk stratification of AML are crucial for guiding treatment decisions, including the use of targeted therapies, the need for hematopoietic stem-cell transplantation, and for predicting patient outcomes.^7–10^ Accordingly, successive updates of the European LeukemiaNet (ELN) recommendations, World Health Organization (WHO) classifications, and International Consensus Classification (ICC) have expanded both the number and types of genetic alterations required for accurate disease classification and prognostication.^4–6,11,12^ AML diagnostics must therefore capture an expanding spectrum of genomic alterations.

Current standard-of-care (SoC) genetic diagnostics for AML rely on a combination of techniques, including chromosome banding analysis (karyotyping), fluorescence in situ hybridization (FISH), targeted polymerase chain reaction (PCR) assays, and targeted next-generation sequencing (NGS) gene panels.^4,6,13^ Although these methods remain the foundation of AML diagnostics, they are constrained by limited resolution or sensitivity, dependence on viable cells, labor-intensive workflows, and, in some cases, reliance on subjective interpretation by clinical laboratory specialists.^13–15^ These limitations highlight the need for a more comprehensive and efficient diagnostic approach.

Whole-genome sequencing (WGS) offers an unbiased, single-assay approach for detecting the broad spectrum of genomic variation, including both protein-coding and non-coding single-nucleotide variants (SNVs), small insertions and deletions (InDels), copy-number alterations (CNAs), and structural variants (SVs).^1,16,17^ Its clinical potential has been demonstrated across multiple malignancies, including lymphoid, pediatric, and solid tumors, where WGS has improved disease classification, revealed cryptic rearrangements, and refined molecular risk stratification.^16–24^ However, few studies have systematically evaluated WGS as a single, comprehensive test for AML classification and risk stratification. In a pioneering study, Duncavage et al. demonstrated that a tumor (leukemia)-only WGS approach could detect recurrent chromosomal rearrangements and mutations in AML-associated genes and facilitate risk stratification according to the ELN 2017 guidelines.^25^ Huber et al. subsequently used tumor-only WGS and whole-transcriptome sequencing in 734 AML cases to assess WHO 2022 and ICC 2022 classification and ELN 2022 risk stratification, without a direct SoC comparison.^26^ However, tumor-only sequencing relies on population databases and prior knowledge to infer the somatic origin of variants, whereas paired tumor-normal sequencing enables direct discrimination of somatic and germline variants, thereby increasing confidence and specificity in the identification of acquired genomic alterations. Despite this advantage, paired tumor-normal WGS has not been systematically benchmarked against contemporary multi-assay SoC diagnostics as a comprehensive diagnostic approach in adult AML under the ELN 2022, ICC 2022, and WHO 2022 classification systems.

As part of an ongoing study within Genomic Medicine Sweden to evaluate WGS as a clinical diagnostic test in acute leukemia^27,28^, we here present a detailed evaluation of the initial retrospective phase, comprising a population-based cohort of 109 adult AML cases. We show that paired tumor-normal WGS can provide a comprehensive diagnostic approach for AML, including detection of clinically relevant genomic alterations, assessment of *TP53* allelic state, and inference of complex karyotype (CK).

## MATERIALS AND METHODS

### Patient data

This study enrolled 109 adult patients diagnosed with AML (including APL) or MDS/AML (as defined by ICC 2022)^5^ admitted to the Department of Hematology at Skåne University Hospital, Lund, Sweden, between 2014 and 2020 (Table 1, Table S1). A subset of patients (n=22; Table S1) is also reported in our multicenter prospective GMS study (Staffas et al., 2026, manuscript submitted to bioRxiv). Throughout this study, “AML” refers collectively to AML and MDS/AML as defined by ICC 2022, unless the distinction is relevant to the analysis or interpretation. For one patient (AML18), only a relapse sample was available and was used for all analyses, including disease classification and ELN 2022 risk stratification. DNA was extracted from bone marrow (n=99) or peripheral blood (n=10). Matched normal DNA was obtained from cultured skin fibroblasts from each patient. This study was approved by the Regional Ethics Committee of Lund University (Dnr 2011/289) and the Swedish Ethical Review Authority (Dnr 2020-06673 and Dnr 2023-01550-01).

**Table 1:** Clinical characteristics of 109 AML cases.

|  |  |  |
|---|---|---|
| Patient characteristics | Number of cases | 109 |
|  | Female sex (number of cases) | 47 |
|  | Median age (years) | 64 |
|  | Median blast percentage in BM (%) | 55 |
| Sample origin<br>(number of cases) | Bone marrow | 99 |
|  | Peripheral blood | 10 |
| Sample timepoint<br>(number of cases) | At diagnosis | 106 |
|  | At relapse | 1 |
|  | During disease | 2 |
| WHO 2022 classification based on SoC<br>(number of cases) | APL with defining genetic abnormalities - *PML*::*RARA* fusion | 6 |
|  | AML with *NPM1* mutation | 41 |
|  | AML with defining genetic abnormalities - other | 22 |
|  | AML, myelodysplasia-related | 24 |
|  | AML defined by differentiation | 6 |
|  | Classified as MDS under WHO 2022 | 10 |
| ICC 2022 classification based on SoC<br>(number of cases) | APL with recurrent genetic abnormalities - *PML*::*RARA* fusion | 6 |
|  | AML with mutated *NPM1* | 41 |
|  | AML with recurrent genetic abnormalities - other | 23 |
|  | AML with mutated *TP53* | 8 |
|  | MDS/AML with mutated *TP53* | 3 |
|  | AML with MR gene mutations | 15 |
|  | MDS/AML with MR gene mutations | 6 |
|  | AML with MR cytogenetic abnormalities | 1 |
|  | AML, not otherwise specified | 6 |
| ELN genetic risk group based on SoC<br>(number of cases) | Acute promyelocytic leukemia with t(15;17)(q22;q21)/*PML–RARA* | 6 |
|  | Favorable risk | 29 |
|  | Intermediate risk | 30 |
|  | Adverse | 44 |
| Antecedent or predisposing conditions<br>(number of cases) | De novo | 83 |
|  | Secondary | 15 |
|  | Therapy-related | 11 |

Abbreviations: AML, acute myeloid leukemia; APL, acute promyelocytic leukemia; BM, bone marrow; WHO 2022, World Health Organization 5th edition of the Classification of Haematolymphoid Tumours; ICC 2022, International Consensus Classification of Myeloid Neoplasms and Acute Leukemias; ELN, European LeukemiaNet; MDS, myelodysplastic syndrome; MR, myelodysplasia-related; NA, data not available

### Standard-of-care diagnostics

All cases included in this study underwent SoC diagnostics, consisting of various combinations of chromosome banding analysis, FISH, targeted PCR/RT-PCR-based assays for recurrent mutation or gene fusion detection, and targeted NGS gene panel analysis, as clinically indicated at the time of diagnosis (Table S2). Since routine SoC gene panel analysis was available only for a subset of cases (n=40),^29,30^ the SoC data were complemented by a rigorously curated whole-exome sequencing (WES) dataset^31^ (mean coverage 95X) for the majority of cases (n=86), including cases lacking gene panel data (n=69). For SNV/InDel comparisons, the SoC comparator therefore comprised available routine molecular testing supplemented by WES to enable more uniform retrospective assessment of small variants across the cohort. Using cytogenetic and molecular results together with blast percentage and clinical history, each case was assigned an SoC-based diagnosis according to the ICC (2022) and WHO (2022) classification systems and a risk group according to ELN (2022).^4–6^

### Whole-genome sequencing

Sample processing and DNA isolation (DNeasy Blood & Tissue Kit, Qiagen, Hilden, Germany) were performed at the Department of Clinical Genetics, Pathology and Molecular Diagnostics, Skåne University Hospital. Library preparation was performed using the Illumina TruSeq PCR-free protocol (Illumina, San Diego, CA, USA), followed by paired-end, 150-bp sequencing on a HiSeq X or NovaSeq 6000 instrument (Illumina). Target coverage depths were 90x for tumor samples and 30x for matched normal tissue samples, and sequencing was performed either at the National Genomics Infrastructure Sweden or at the Department of Clinical Genetics, Pathology and Molecular Diagnostics.

### Bioinformatic analysis and variant matching

Two bioinformatic pipelines were used to analyze the tumor-normal paired WGS data: 1) DRAGEN secondary analysis (herein referred to as WGS-DRAGEN) (Illumina Inc; hardware version 07.021.624, software version 3.10.4), and 2) nextflow_tumwgs (herein referred to as WGS-SMD),^32^ an in-house-developed pipeline at the Section of Molecular Diagnostics, Skåne University Hospital. This parallel analysis enabled comparison of pipeline performance and provided complementary information that strengthened variant interpretation. Pipeline-specific performance is only reported where informative.

Alignment to GRCh38 and read deduplication were performed within each pipeline. Panels of normal variants were constructed for WGS-DRAGEN (based on 20 normal samples) and WGS-SMD (50 normal samples). WGS-SMD used FreeBayes^33^ (v1.3.1) and VarDict^34^ (v1.7.0) for SNV/InDel detection, Manta^35^ (v1.6.0) for SV detection, and a custom GATK-based^36^ (v4.1.4) implementation for CNA detection. WGS-DRAGEN used custom algorithms for detection of SNVs, SVs, and CNAs. All variants were annotated with Ensembl VEP^37^ (release 103). The variants were further filtered uniformly using caller-derived quality scores and PASS status; for SVs, split and spanning read support and breakpoint quality were also considered, with more lenient criteria for known recurrent SVs.

SNV/InDel variants with a variant allele frequency (VAF) ≥5% in at least one of the SoC, WGS-DRAGEN, or WGS-SMD pipelines and a VEP-predicted impact of moderate or higher were included. Variant calls with a VAF <5% were also included if another pipeline reported the same variant at ≥5% VAF. Variant liftover (for gene panel and WES variants in the hg19 reference), normalization, and matching between WGS and SoC pipelines were performed using Picard (v2.22.1; https://github.com/broadinstitute/picard), BCFtools^38^ (v1.11) and a custom Python script. A total of 19 variants were matched manually by consolidating multi-event calls into unified variants (n=12) and by resolving mismatched alternate-allele descriptions for *FLT3*-ITD detection in SoC and WGS workflows (n=7) (Table S3). The *in-silico* gene list used to compare WGS results with SoC was defined by either the GMS Myeloid Gene Panel,^30^ comprising 191 genes (90 cases) or the TruSight Myeloid Sequencing Panel (Illumina), including 54 genes (19 cases; Table S2). The overlap between these two gene lists was 53 of 54 genes (with *CBLC* not present in the GMS Myeloid Gene Panel).

For SoC–WGS SNV/InDel variant comparisons, a set of true-positive reference variants was defined as the union of SoC-reported variants and high-confidence variants detected by WGS (variants called by both WGS-DRAGEN and WGS-SMD, with a predicted impact of moderate or higher). Variants affecting *U2AF1* were included despite being present in only one of the WGS pipelines since a known issue affecting regions of high sequence homology in the GRCh38 reference prevented detection by DRAGEN.^39^ This has since been corrected in DRAGEN version 4.5 (Illumina, personal communication). Consequently, WGS-only variants were identified by both WGS pipelines but not SoC, SoC-only variants were detected by SoC and at most one of the WGS pipelines, and fully concordant variants required detection by SoC and both WGS pipelines. All WGS-only and SoC-only variants were manually reviewed to confirm their presence or absence in both WGS and SoC data.

### Cytogenetic abnormalities and complex karyotype

The comparison of cytogenetic abnormalities detected by SoC and WGS included all aberrations relevant for any of the three classification systems (ELN, ICC, and WHO). These were grouped into the following categories: −17/abn(17p), −5/abn(5q), −7/del(7q), +8, del(11q), abn(12p), - 20/del(20q), and idic(X)(q13). Here, abn() denotes any gain or loss of chromosomal material larger than 3 Mbp, as well as structural events such as inversions, translocations, or insertions. WGS-based analysis of the *TP53* locus specifically included events predicted to cause transcript ablation, deletions, and copy-neutral loss-of-heterozygosity (CN-LOH).

When describing the genetic findings in individual cases, CK based on WGS was defined as three or more unrelated chromosomal abnormalities larger than 3 Mbp, without considering the presence of class-defining recurrent genetic abnormalities (RGAs). CK by SoC was defined as three or more unrelated chromosomal abnormalities detected by conventional cytogenetics in at least three metaphases, also irrespective of the presence of RGAs. However, for classification and risk stratification, CK was assigned (from either WGS or SoC) only in the absence of higher-priority RGAs, in accordance with current classification hierarchies.

To estimate CK from WGS data in an objective, automated, and reproducible manner, we developed the CK probability score (CK-prob), a supervised machine-learning logistic regression model that was subsequently applied uniformly across the cohort. The model was trained and evaluated based on SoC-defined CK status (including cases with RGAs). The following predictors from WGS data were included in the final model: the fraction of the genome affected by CNA, weighted by copy number and scaled to 100 (wFGA100); the number of high-confidence SVs (nSV); and the number of chromosomes affected either by a substantial CNA (at least 5% of chromosome length) or by a high-confidence SV (nChr). Predictors were normalized using a z-score transformation to ensure comparable scaling.

Model fitting was performed using L2-regularized logistic regression implemented in scikit-learn^40^ (v1.8.0). Model discrimination was evaluated using stratified 10-fold cross-validation, with the area under the receiver operating characteristic curve (AUC) calculated from pooled out-of-fold predicted probabilities. Nonparametric bootstrap resampling (5,000 iterations) was used to estimate 95% confidence intervals (CIs). For regression coefficients and corresponding odds ratios (ORs), the model was refitted in each bootstrap sample, including feature scaling. The resulting model achieved a cross-validated AUC of 0.875 (95% CI 0.758–0.970). For direct application to unstandardized predictor values, the coefficients and intercept of the final model were transformed back to the original predictor scale, yielding the following CK-prob equation:

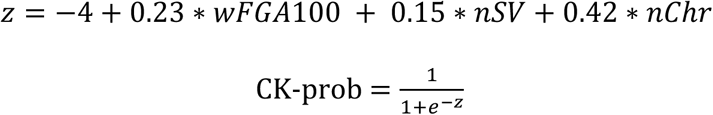

### Statistical analyses

Sensitivity, precision, and F1-scores were calculated against the true-positive reference variant set using standard formulas. SciPy^41^ (version1.14.1) was used to calculate correlation coefficients and perform statistical tests, including t-tests and Mann-Whitney U-tests.

## RESULTS

### Standard-of-care and WGS results

To evaluate WGS as an integrated diagnostic approach in AML, we performed short-read sequencing of paired tumor-normal samples representing 109 AML cases and compared the results with SoC diagnostics (Figure 1, Table 1, Tables S1 and S2). WGS generated high-quality data for all specimens, with a mean coverage of 108x for leukemia samples and 34X for matched normal samples. In contrast, G-band karyotyping failed in two cases (Table S1).

**Figure 1.**
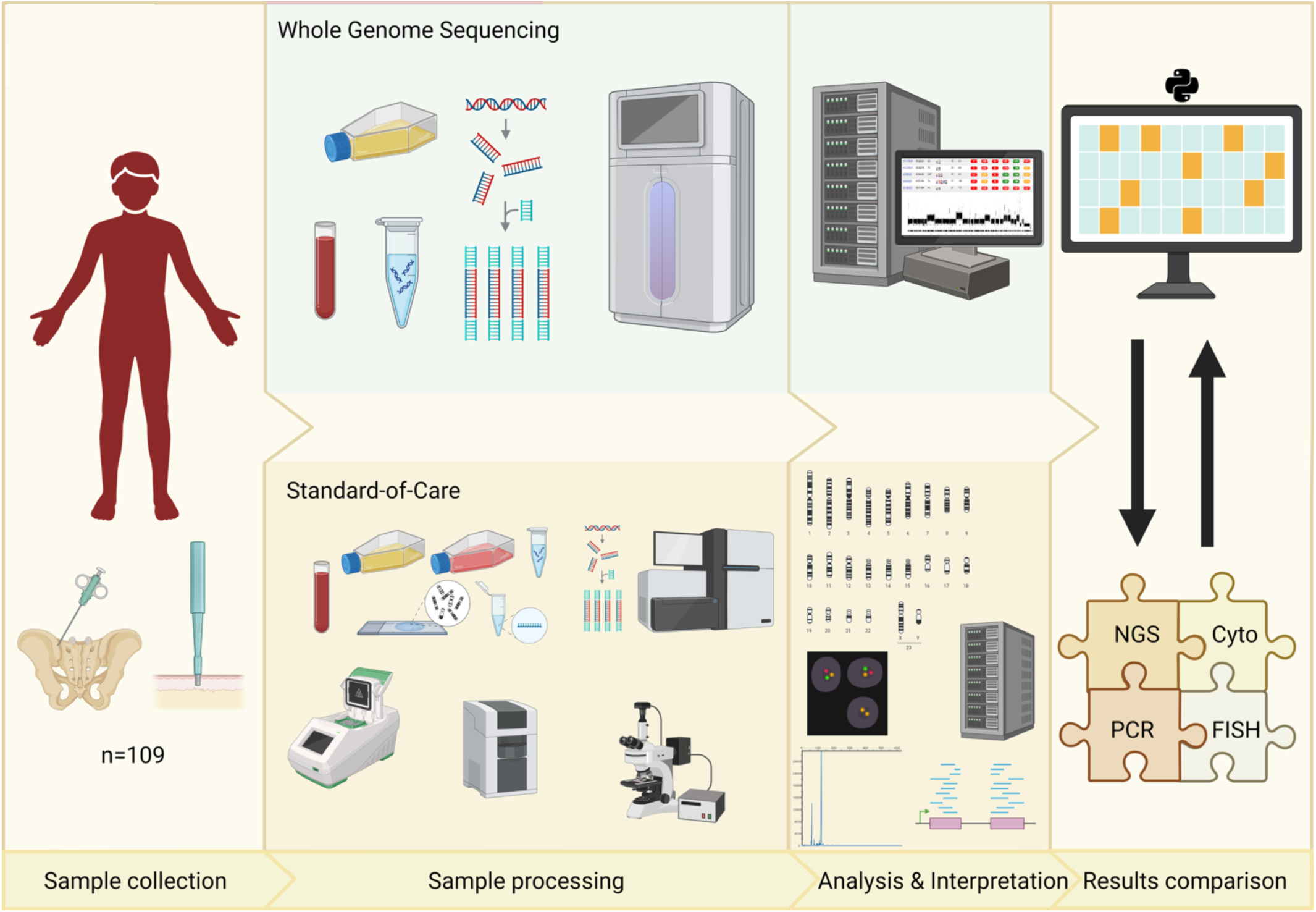
Study design and diagnostic workflow. Schematic overview of the study workflow from sample collection to results comparison. Paired leukemia and cultured skin-fibroblast samples from 109 adult patients underwent whole-genome sequencing (WGS; upper flowchart) followed by bioinformatic analysis. The standard-of-care (SoC; bottom flowchart) workflow included cytogenetics (Cyto), fluorescence in situ hybridization (FISH), targeted polymerase chain reaction (PCR), fragment analysis (FA), and next-generation sequencing (NGS) assays. Data from WGS and SoC were analyzed independently. The results were then compared with regard to detected variants and the effect on disease classification, risk stratification, and the estimated genomic complexity.

### WGS shows high concordance with standard-of-care analyses and improves small-variant detection

A total of 443 SNVs/InDels were detected by either WGS or SoC analyses within 191 clinically relevant genes (Figure 2A, Table S3). Of these, 395 (395/443; 89%) variants were detected by both SoC and WGS, highlighting high concordance between the analyses. Thirteen variants (13/443; 3%) did not meet the predefined technical criteria for WGS detection and were therefore classified as SoC-only (Figure 2A). Three of these (3/13; 23%) were detected by WGS-DRAGEN only (but not the other WGS pipeline as required for WGS detection), including an *NPM1* mutation in AML63. Eight variants (8/13; 62%) were represented by few reads in WGS data but were not called by either pipeline. No supporting WGS reads were identified for the remaining two variants (2/13; 15%), both of which had low VAFs in the SoC/WES analysis (5% and 6% for *KMT2C* and *GFI1*, respectively). Thus, the presence of small subclones combined with methodological differences (PCR-free preparation for WGS) and stochastic sampling at ∼90x WGS coverage may explain the discrepancies.

**Figure 2.**
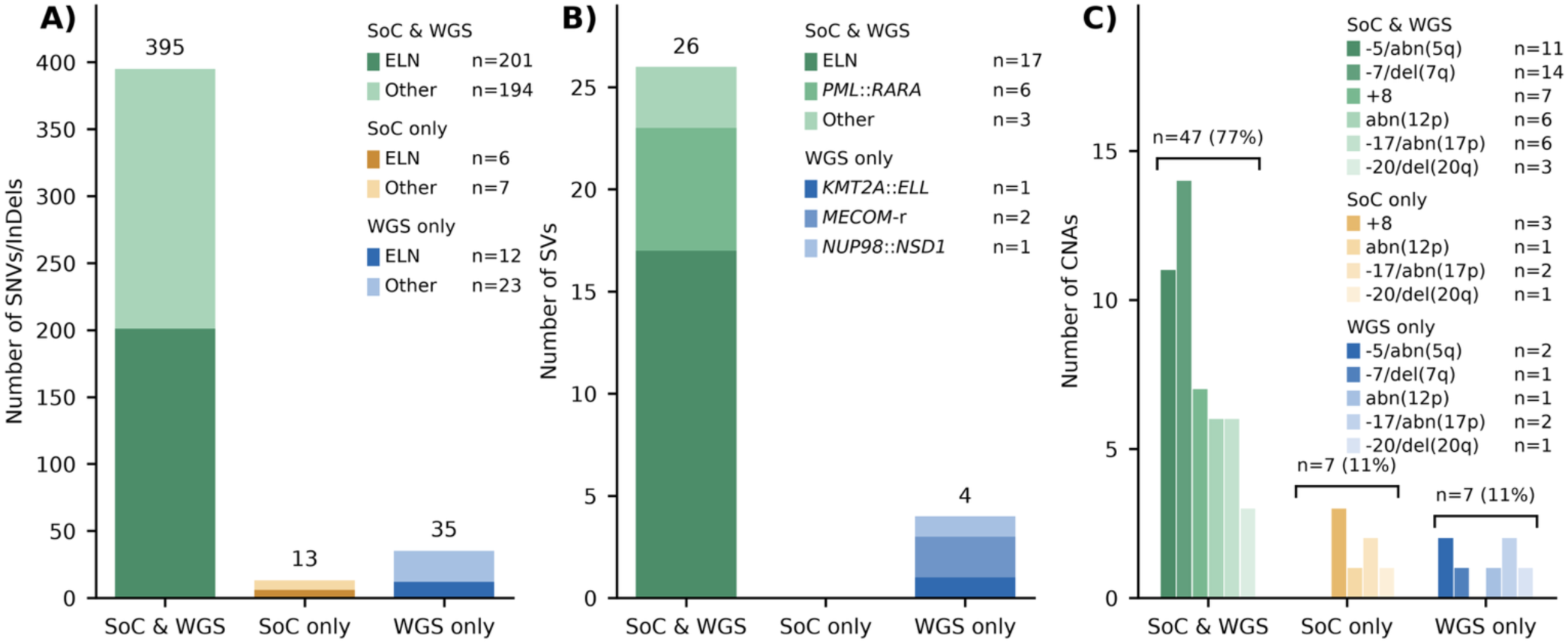
Concordance of variant detection between standard-of-care diagnostics and whole-genome sequencing. Variants detected by both standard-of-care (SoC) diagnostics and whole-genome sequencing (WGS) are shown in green, SoC-only variants in yellow, and WGS-only variants in blue. Detection categories follow the criteria described in Methods. **(A**) Single-nucleotide variants (SNVs) and small insertions/deletions (InDels), with darker shades indicating variants in genes used for European LeukemiaNet (ELN) 2022 risk stratification with addition of *IDH1* and *IDH2*. **(B)** Structural variants recurrent in AML. Concordant events are grouped as ELN-defined risk-associated rearrangements, *PML::RARA*, or other recurrent rearrangements; WGS-only variants are grouped by rearrangement type. No structural variants were detected exclusively by SoC. **(C)** Copy-number alterations relevant to ELN 2022 risk stratification or WHO/ICC 2022 classification, grouped by affected chromosome or region. Percentages indicate the proportion of all alterations included in panel C.

Conversely, 35 variants (35/443; 8%) were detected by WGS only. Of these, approximately half (n=18) had no or low coverage in SoC data, including a splice donor variant in *STAG2* with low WES coverage (14X). Fifteen WGS-only variants had supporting reads in the SoC analysis, but were below the limit of detection, as the majority had either low coverage (<30x), low theoretical VAF (<3%) based on the SoC reads, or both. These included two c.1934dupG mutations (38% and 41% VAFs) in the clinically significant *ASXL1* gene. Detection of this mutation, occurring within a mononucleotide guanine repeat, is known to be influenced by replication slippage during PCR.^42^ Thus, the PCR-free WGS protocol appears to improve the detection accuracy of *ASXL1* c.1934dupG. Lastly, clinically important *FLT3*-ITD mutations were detected by WGS in two AMLs (AML24 and AML72), despite being negative in the targeted SoC *FLT3*-ITD assays. These unexpected WGS findings prompted renewed analyses using the SoC PCR method, which confirmed that both samples were positive, in line with the WGS data. Since the original SoC analysis had been performed at an external laboratory, the exact reason for the false-negative results could not be investigated further.

VAFs correlated strongly between SoC and WGS (Pearson r = 0.873 for WGS-DRAGEN; Figure S1). Overall, the sensitivity of SNV/InDel detection was significantly higher for WGS than for SoC: 97.1% (95% CI, 95.5%–98.6%) versus 92.1% (95% CI, 89.6%–94.6%) for SoC. WGS-DRAGEN alone reached 96.6% sensitivity (95% CI, 94.8%–98.2%), whereas WGS-SMD reached 97.1% sensitivity (95% CI, 95.5%–98.6%). However, between the two WGS pipelines, WGS-DRAGEN reported markedly fewer potentially false-positive findings (n=2) compared with WGS-SMD (n=104; Figure S2, Table S4). This reflects the design of WGS-SMD, which favors sensitivity over specificity and relies on a final manual review of variants. F1 scores were higher for WGS (98.5%; CI 95%, 97.7%-99.3%) compared to SoC (95.9%; CI 95%, 94.5%-97.2%) and WGS-DRAGEN (98.1%; CI 95%, 97.1%-99%) compared to WGS-SMD (88%; CI 95%, 86%-90%).

### WGS improves detection of clinically relevant structural variants

We next compared detection of guideline-relevant recurrent SVs between WGS and SoC. In total, 30 SVs were detected by either method (Figure 2B; Table S5), of which 26 (26/30; 87%) were concordant. No SVs were detected by SoC only, whereas four (4/30; 13%) were identified only by WGS. Two of these involved *MECOM*: a *GATA2*::*MECOM* rearrangement consistent with inv(3)(q21q26) in AML183 and a *MECOM*::*MYC* representing t(3;8) in AML55, both missed by cytogenetics. Increased *MECOM* expression was confirmed using previously published RNA-seq data^31^ from these two cases (Figure S3). The remaining WGS-only SVs were a cytogenetically cryptic *KMT2A*::*ELL* fusion in AML336, corresponding to t(11;19), and a *NUP98*::*NSD1* rearrangement in AML237, resulting from t(5;11)(q35;p15). Importantly, the detection of *KMT2A*::*ELL* in AML336 altered not only WHO and ICC classifications but also changed the ELN risk category from intermediate to adverse. Manual interpretation of WGS data was required to detect a *GATA2::MECOM* in AML343. A tandem duplication and large deletion were indicated, juxtaposing *MECOM* with the *GATA2* distal enhancer on 3q21. This combined result was not noticed by the variant effect prediction software. The detected breakpoints were consistent with a t(3;3)(q21;q26) as reported by cytogenetics.

### WGS improves detection of clinically relevant copy-number alterations

We next compared the ability of WGS and SoC to detect guideline-relevant CNAs (Figure 2C). A total of 61 such CNAs were identified by either SoC or WGS. Of these, 47 (77%) were detected by both methods, demonstrating overall good concordance across platforms. Concordant events included most −5/abn(5q), −7/del(7q), trisomy 8, abn(12p), -17/abn(17p), and -20/del(20q) (Figure 2C; Table S6).

Seven CNA events were detected by SoC only, including trisomy 8 (n=3), abn(12p) (n=1), abn(17p) (n=2) and monosomy 20 (n=1). Aneuploidies of chromosomes 8 and 20 were subclonal events in all four SoC-only cases. Trisomy 8 was called by one WGS pipeline (WGS-DRAGEN) in one case (AML54), and was undetected by either WGS pipeline in the remaining two cases (AML52 and AML78). In AML119, SoC analysis described an add(17)(p11) while WGS detected a 598 kbp deletion on 17p that included the *TP53* locus. These were too dissimilar to be considered concordant. In contrast, seven events were detected by WGS only: two −5/abn(5q), one −7/del(7q) alterations, one abn(12p), two 17p abnormalities, and one del(20q). WGS-only findings included a relatively small (4.63 Mbp) distal 5q gain in AML175 and focal 17p losses in AML62, consistent with the limited resolution of chromosome banding. Other discrepancies involved larger losses in cases with complex karyotypes (AML44, AML51, AML83) or limited number of representative metaphases (AML56).

### WGS detects focal structural and copy-number alterations affecting AML-associated genes

Beyond these guideline-relevant alterations, WGS can detect medium-sized structural variants and focal CNAs that are typically not identified using SoC methods (ranging from 150 bp to 300 kbp; excluding *FLT3*-ITD). A number of such intragenic SVs and CNAs have been described in AML-associated genes,^43,44^ but they are currently not used for AML subtype classification. In our dataset, four patients with *KMT2A* partial tandem duplications (*KMT2A*-PTD) were identified (Figure 3; Table S7). In *RUNX1*, one case with a focal deletion, one with tandem duplication, and two with intragenic insertions were detected, demonstrating multiple mechanisms for gene truncation.

**Figure 3.**
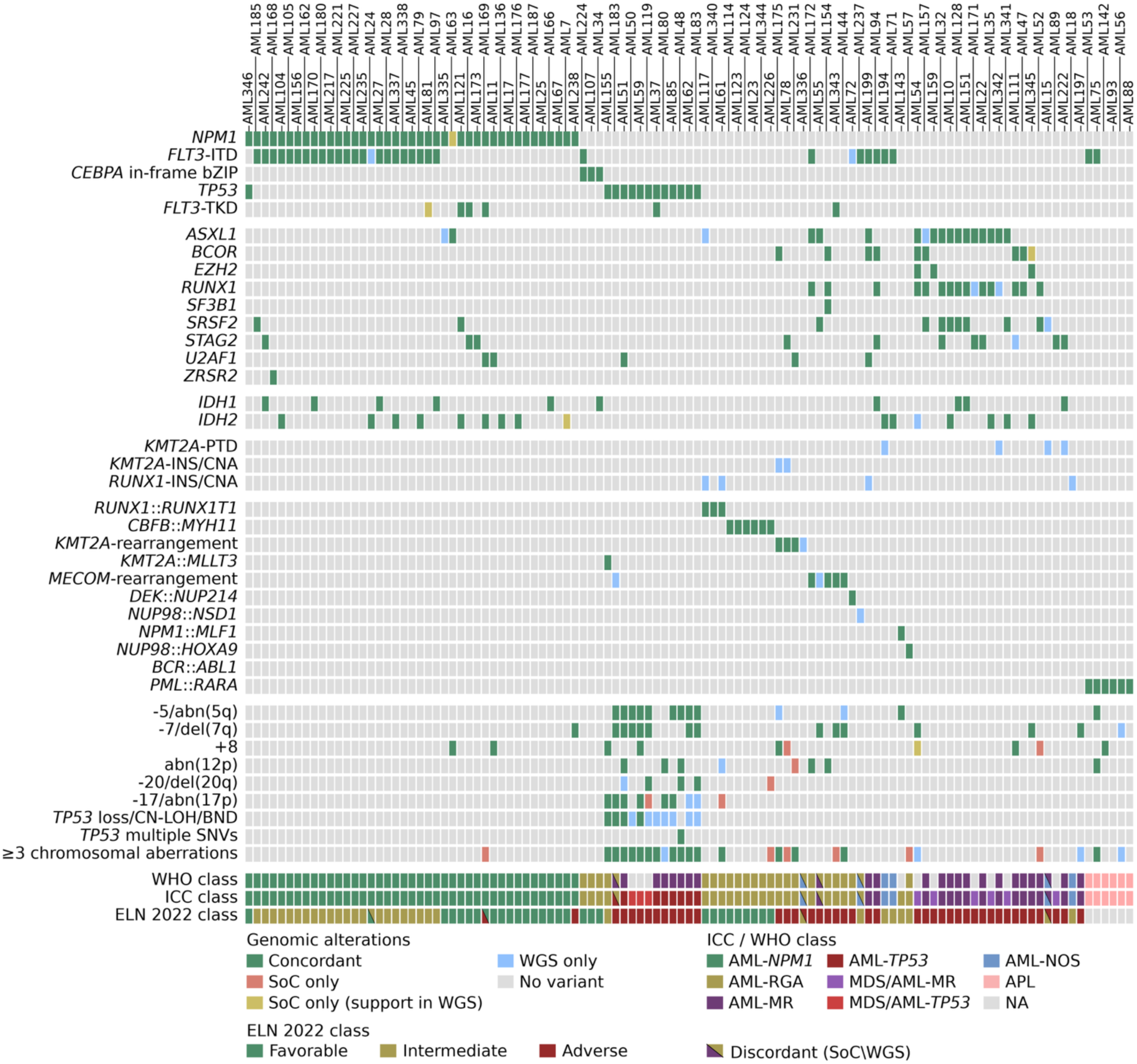
Genomic alterations and diagnostic concordance across the AML cohort. Tile plot showing selected genomic alterations, International Consensus Classification (ICC) 2022, World Health Organization (WHO) 2022 and European LeukemiaNet (ELN) 2022 risk assignments (in rows) for 109 cases (in columns). For genetic-event rows, green indicates concordant detection, blue WGS-only detection, red SoC-only detection without supporting WGS evidence, and yellow SoC-reported alterations supported by manual WGS data examination but not meeting the predefined WGS detection criteria. In the risk stratification and classification rows, the full rectangles mark agreement between SoC and WGS, whereas diagonally divided rectangles show the change between SoC class (bottom-left) and WGS (top-right). AML-RGA represents AML with recurrent genetic abnormalities (ICC) or Acute myeloid leukaemia with defining genetic abnormalities (WHO) excluding AML with *NPM1* mutation and APL *PML::RARA*, which are displayed separately. Abbreviations: APL, acute promyelocytic leukemia; BND, breakend; bZIP, basic leucine zipper; CNA, copy-number alteration; CN-LOH, copy-neutral loss of heterozygosity; INS, insertion; ITD, internal tandem duplication; MDS, myelodysplastic neoplasm; MR, myelodysplasia-related, encompassing gene mutations and cytogenetic abnormalities; NA, not applicable or not classified; NOS, not otherwise specified; PTD, partial tandem duplication; RGA, recurrent genetic abnormalities; SNV, single-nucleotide variant; TKD, tyrosine kinase domain.

### WGS-based classification and risk stratification show high concordance with SoC across ELN, ICC, and WHO 2022 frameworks

Having established high concordance and improved detection across major variant classes, we next compared subtype classification and risk-stratification based independently on data from SoC or WGS (Figure 3). ELN 2022 risk group was established in 103 eligible cases (excluding six APLs but including MDS/AML) using either SoC or WGS data. SoC assigned an ELN risk category based on qualifying genetic findings in 99 cases, while four cases (AML15, AML18, AML57, AML336) lacked qualifying alterations and were therefore classified to intermediate risk. Using the WGS-based workflow, 101 of 103 cases had qualifying genetic findings for ELN risk assignment, with no relevant abnormalities detected in AML18 and AML57 (Figure 3). Overall, ELN risk assignment was highly concordant between SoC-and WGS-based classifications, with only four discrepancies (4/103; 4%; Figure 4A; Tables S1 and S8). AML15 was reclassified from intermediate to adverse risk due to detection of an *SRSF2* mutation by WGS. AML24 moved from favorable to intermediate risk due to a *FLT3*-ITD detected only by WGS. AML169 with *NPM1* mutation was reclassified from adverse to favorable risk because the SoC-based complex karyotype (CK) designation was not supported by WGS. Finally, AML336 was reclassified to the adverse risk group based on the detection of a *KMT2A*::*ELL* fusion by WGS.

**Figure 4.**
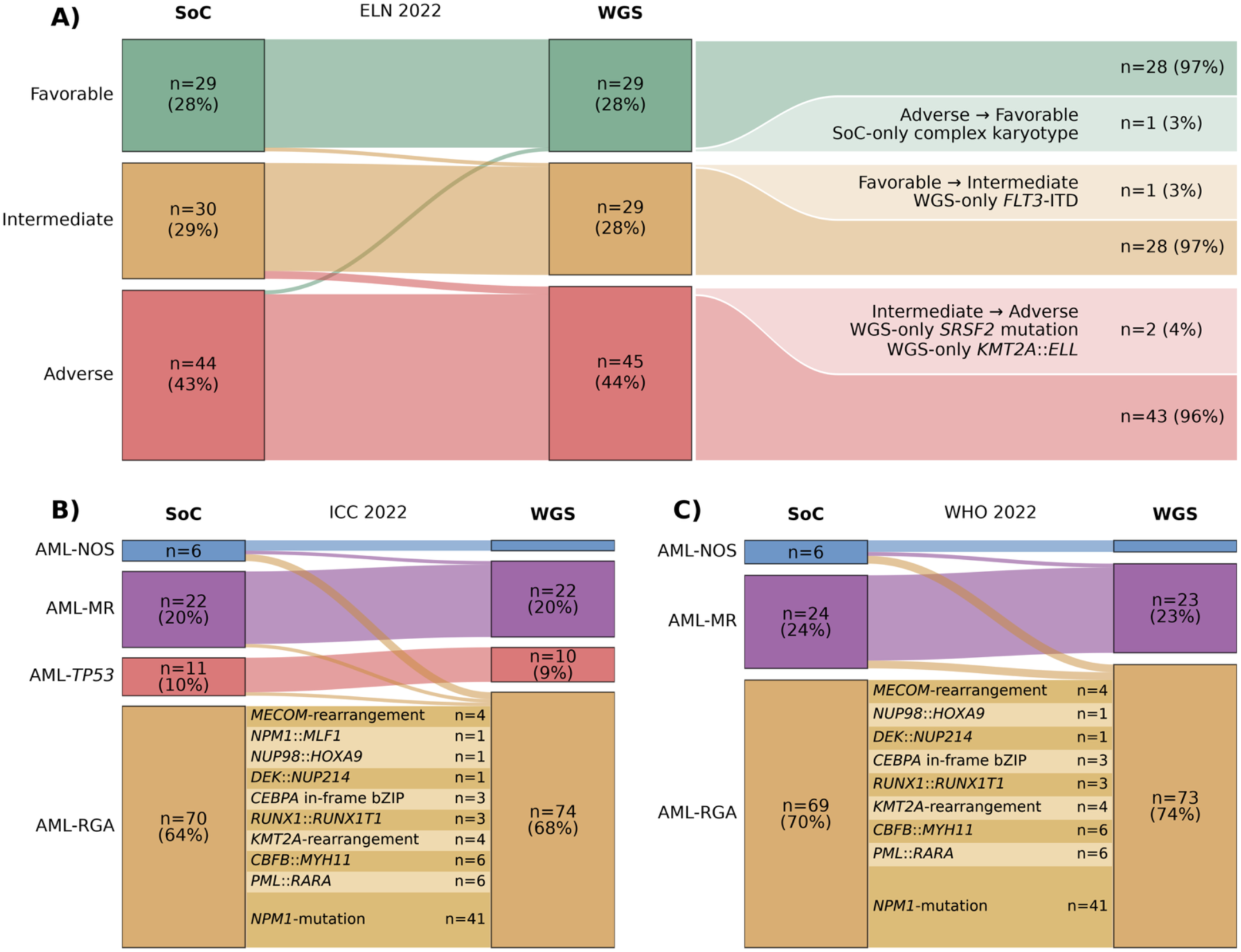
Changes in ELN, ICC, and WHO 2022 classifications based on standard-of-care versus whole-genome sequencing. Alluvial plots comparing risk group and subtype assignments based on standard-of-care (SoC; left) and whole-genome sequencing (WGS; right) data. MDS/AML was not treated as a separate entity. Such cases were included under AML groups in the ELN and ICC plots (but excluded from the WHO plot). **(A)** ELN 2022 risk stratification of 103 eligible cases, excluding six cases of acute promyelocytic leukemia. Risk assignments were concordant in 99/103 cases (96%) while four cases changed risk category based on novel WGS findings. **(B)** International Consensus Classification (ICC) 2022 classification of 109 cases, including MDS/AML, displayed in grouped categories. AML-MR group includes both AML with myelodysplasia-related gene mutations (n=21) and AML with myelodysplasia-related cytogenetic abnormalities (n=1); AML-RGA encompasses class-defining recurrent genetic abnormalities. Assignments were concordant in 104/109 cases (95%). **(C)** World Health Organization (WHO) 2022 assignments in 99 eligible cases (excluding MDS/AML), with concordance in 94/99 cases (95%). In both **(B)** and **(C)**, five reclassifications followed WGS-only detection of an *SRSF2* mutation, *NUP98*::*NSD1*, *KMT2A*::*ELL*, and two *MECOM* rearrangements. AML-NOS denotes AML not otherwise specified in the ICC and, for display purposes, AML defined by differentiation in the WHO classification.

Two cases lacked ELN-defining alterations by both approaches. AML18 harbored mutations in *ARID2*, *USP9X*, *BCORL1*, and *GATA2*, none of which are included in ELN 2022. In this case, a *RUNX1* tandem duplication of unknown significance involving exon 5 was detected by WGS only. AML57 carried *JAK2* and *TET2* mutations and a *NUP98::HOXA9* fusion, none of which affect ELN 2022 risk assignment. In addition, WGS revealed a novel *CREB5::WDR88* fusion in AML197, complementing monosomy 7 identified by both WGS and SoC.

We next assessed AML and MDS/AML classification according to ICC 2022 and WHO 2022 using SoC and WGS independently. Concordance remained high, with 95% agreement in both ICC and WHO (Figure 4B–C). Using subtype classification based on ICC 2022, five cases (5/109, 5%) were reclassified based on WGS findings. AML15 shifted from AML-NOS (AML, not otherwise specified) to AML-MR (AML with myelodysplasia-related gene mutations) due to the detection of an *SRSF2* mutation. AML237, AML336, AML183, and AML55 were reclassified as AML with RGAs following detection of *NUP98*::*NSD1*, *KMT2A::ELL* and two *MECOM*-rearrangements respectively. The same discordances described above also resulted in five corresponding reclassifications based on WHO 2022.

### WGS refines *TP53* allelic state determination and assessment of complex karyotype

Both *TP53* mutations and 17p aberrations, typically affecting the *TP53* locus, are associated with adverse prognosis and CK in AML.^45,46^ We therefore examined whether integrated WGS analysis of SNVs, CNAs, and SVs could resolve *TP53* allelic state and support WGS-based assessment of CK.

*TP53* SNV status was fully concordant between SoC and WGS, with 13 cases harboring *TP53* mutations (Figure 5A, Table S3). By integrating SNV, CNA, SV and CN-LOH data, WGS identified a second event consistent with biallelic *TP53* disruption (bi*TP53*) in 12 of the 13 cases (12/13; 92%). All bi*TP53* AMLs harbored more than three unrelated chromosomal aberrations as determined by WGS. Eleven of these twelve cases were classified as CK by SoC. In addition, only seven (7/12; 58%) bi*TP53* AMLs displayed a VAF above 50%, and only eight (8/12; 67%) showed evidence of a second-hit event involving *TP53* by SoC (Figure 5A). This indicated that bi*TP53* status was incompletely resolved by SoC in a substantial proportion of cases. The sole *TP53*-mutated case without bi*TP53* (AML346) harbored an *NPM1* mutation and lacked CK, suggesting a different genetic background of this case (Figure 5A, Table S9).

**Figure 5.**
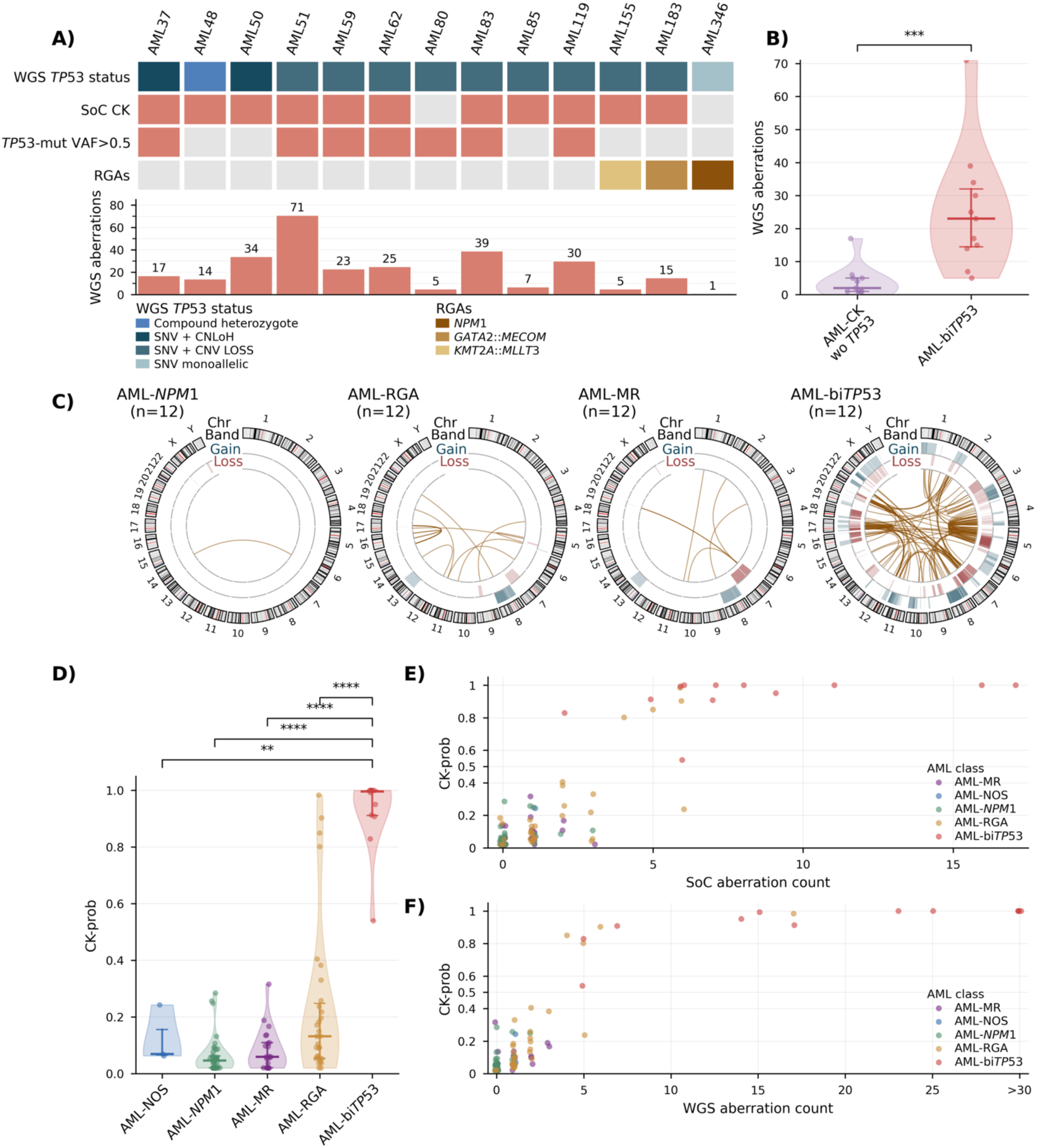
Integrated assessment of *TP53* allelic state and genomic complexity by WGS. **(A)** Whole-genome sequencing (WGS)-inferred *TP53* allelic status, standard-of-care (SoC) complex karyotype (CK), *TP53* variant allele frequency (VAF) >0.5, and co-occurring recurrent genetic abnormalities (RGAs) in the 13 *TP53*-mutated cases. Salmon and gray tiles indicate the presence and absence, respectively, of SoC CK or VAF >0.5. Bars show the number of WGS-defined chromosomal aberrations per case. **(B)** Violin plot showing the distribution of WGS-defined aberration counts in *TP53*-wildtype cases with CK by SoC (AML-CK wo *TP53*) and in cases with biallelic *TP53* alterations (AML-bi*TP53*). Points represent individual cases; statistical significance was assessed using a two-sided Mann–Whitney U test (*** P < 0.001). (**C)** Circos plots summarizing alterations in 12 randomly selected cases from each of the AML–*NPM1*, AML-RGA, and AML-MR groups, and all 12 AML-bi*TP53* cases. Tracks show chromosome ideograms and copy-number gains in blue and losses in red, with increasing opacity with the number of alterations. Links represent structural variants. **(D)** Distribution of CK-prob across AML subgroups: AML with myelodysplasia related gene mutations or cytogenetic abnormality (AML-MR; purple), AML not otherwise specified (AML-NOS; blue), AML with mutated *NPM1* (AML-*NPM1*; green), AML with recurrent genetic abnormalities excluding *NPM1* mutations (AML-RGA; yellow), and biallelic *TP53* alterations (bi*TP53*, including cases with bi*TP53* and RGA; red). Violin plots with overlaid data points illustrate group-wise distributions. Statistical comparisons were performed using two-sided Mann–Whitney U tests; significance levels are indicated as P <0.05 (*), <0.01 (**), <0.001 (***), and <0.0001 (****). For subgroup comparisons, AML-bi*TP53* cases are grouped together irrespective of co-occurring class-defining abnormalities, and AML–*NPM1* is shown separately from other AML-RGA. **(E)** Scatter plot showing the relationship between number of unrelated aberrations as detected by SoC cytogenetics and WGS-based CK-probability score (CK-prob) across AML subgroups. **(F)** Scatter plot showing the relationship between number of aberrations as detected by WGS (after data curation) and CK-prob score across AML subgroups. WGS aberration counts >30 are displayed at a common x-coordinate.

Cases with bi*TP53* showed substantially greater genomic complexity than other AMLs, with more WGS-detected SVs and CNAs (Figures 5B-C, Figures S4-S10), supporting the view that CK in AML largely serves as a surrogate marker of bi*TP53*.^45,47–49^ Compared with SoC-determined CK cases lacking a *TP53* mutation, bi*TP53* cases had markedly more chromosomal aberrations by WGS (median 20 vs 3; p<0.001, Mann-Whitney U-test; Figure 5B).

To automate and standardize WGS-based CK assessment, we developed the CK-probability score (CK-prob), a supervised machine-learning model trained against SoC-defined CK. The final model incorporated three WGS-derived metrics: the fraction of the genome affected by CNAs, SV burden, and the number of chromosomes affected by substantial CNAs or SVs (Table 2, Figure S11). At a threshold of 0.5, CK-prob classified 16 AMLs as CK-positive, including all twelve bi*TP53* cases and four additional AML-RGA cases (Figure 5D, Table S10). The four additional CK-prob-positive cases harbored class-defining RGAs that take precedence over CK in disease classification and at least four unrelated chromosomal aberrations: AML44 (*MECOM* rearrangement; CK-prob 0.98), AML175 (*KMT2A* rearrangement; 0.90), AML61 (*RUNX1*::*RUNX1T1*; 0.85), and AML75 (*PML*::*RARA*; 0.80).

**Table 2:** CK-prob regression model description.

|  | Coefficient value (scaled) | Coefficient value (original) [95% CI] | Odds ratio [95% CI] |
|---|---|---|---|
| α (intercept) | -1.5 | -4 [-6.2; -2.9] |  |
| β<sub>1</sub> (*wFGA100* coef) | 1.25 | 0.23 [0.08; 0.44] | 1.26 [1.08; 1.55] |
| β<sub>2</sub> (*nSV* coef) | 1.3 | 0.15 [0.02; 0.5] | 1.17 [1.02; 1.66] |
| β<sub>3</sub> (*nChr* coef) | 1.3 | 0.42 [-0.03; 0.91] | 1.52 [0.97; 2.49] |

Abbreviations: wFGA100, weighted fraction of the genome affected by CNAs, scaled to 100; nSV, number of structural variants; nChr, number of unique chromosomes involved in either substantial CNAs or SVs

We compared CK-prob with aberration counts obtained by SoC cytogenetics and manual WGS aberration counting. SoC identified at least three unrelated aberrations in 22 cases, including 11 of the 12 bi*TP53* AMLs and 11 cases assigned to other AML classes (9 AML-RGA, 1 AML-MR, and 1 AML-*NPM1*; Figure 5E, Table S10). At the same threshold, manual WGS counting identified 20 cases, including all 12 bi*TP53* AMLs and eight additional cases (6 AML-RGA and 2 AML-MR). Raising the WGS threshold to five aberrations retained all 12 bi*TP53* AMLs and four additional cases, all AML-RGA (Figure 5F, Table S10).

Collectively, integrated WGS analysis refined *TP53* allelic state determination and enabled automated assessment of genomic complexity.

## DISCUSSION

Contemporary AML diagnostics require simultaneous assessment of gene mutations, SVs, CNAs, and karyotypic complexity to assign patients according to the most recent classification and risk stratification schemes.^4–6^ In a population-based cohort of 109 adult AML cases, we here show that paired tumor-normal WGS achieves high concordance with current SoC diagnostics, including 95% concordance for WHO 2022 and ICC 2022 classifications, and 96% for ELN 2022 risk assignment, while identifying clinically relevant alterations missed by conventional workflows, leading to reclassification in 4–5% of cases. We also demonstrate that WGS improves *TP53* allelic state determination compared with SoC and introduce a novel WGS-derived scoring algorithm, the CK-prob score, to automate CK identification from WGS data.

WGS has previously compared favorably against conventional cytogenetics and targeted sequencing in both pediatric/adolescent ^21^ and adult^25^ AML cohorts. Duncavage et al.^25^ demonstrated improved ELN 2017 risk assignment in 14.7% of AML cases using tumor-only WGS, while Voss et al.^21^ showed improved detection of cryptic rearrangements using integrated WGS/WTS in 153 pediatric and adolescent AML cases. Recently, Huber et al. ^26^ applied tumor-only WGS and WTS to a large adult AML cohort to examine the consequences of the WHO 2022, ICC 2022, and ELN 2022 frameworks, but without directly benchmarking WGS against a multi-assay SoC diagnostic workflow. Building on these studies, we evaluated paired tumor-normal WGS against a contemporary multi-assay SoC workflow and assessed its diagnostic consequences under current 2022 WHO, ICC, and ELN guidelines. This analysis showed near-complete concordance, with only a small number of reclassifications resulting mainly from improved SV detection or reinterpretation of genomic complexity by WGS (4/103, 5/109, and 5/99 cases were reclassified using the ELN, ICC and WHO systems, respectively). Accordingly, the improved diagnostic yield observed using WGS with previous classification systems^25,50^ remains evident under the current more mutation-focused frameworks.

The detection of SNVs and InDels was highly concordant between SoC and WGS. The few discrepancies observed involved regions not covered by targeted methods, regions affected by PCR artifacts, and variants with low VAF. Thus, the 90x sequencing depth WGS employed here allowed reliable detection of variants at VAFs around 5%, in accordance with previous studies with 60x-90x coverage. ^21,25^ All SNV and InDel variants detected by SoC but not detected by WGS had low VAF by SoC, and even lower support by WGS reads, indicating that they represent variants in minor subclones that could potentially also be detected by WGS with increased sequencing depth. Conversely, among the WGS-only variants, there were 12 (11%) SNVs/InDels in ELN-included genes, including four cases with *ASXL1* alterations and two cases with *FLT3*-ITD. The latter represented false-negative findings at an external laboratory that could not be scrutinized further. Nevertheless, this highlights another advantage of WGS as a unified assay, where fewer individual analyses leave less room for human errors. Comparison of two independent bioinformatic pipelines—a commercial platform (WGS-DRAGEN) and an in-house solution (WGS-SMD)—revealed how pipeline design and tuning can affect variant detection and reporting. Although both showed high sensitivity for SNVs/InDels (97.7% and 97.1%, respectively), the differences in precision and reporting indicate that clinical implementation of WGS requires systematic pipeline validation and awareness of pipeline-specific limitations.

For SVs and CNAs, WGS provided additional information beyond conventional cytogenetics and improved diagnostic yield, in line with recent studies in adult and pediatric AML.^21,25^ WGS provided genome-wide chromosomal assessment in all cases, including the two without an informative SoC karyotype. This is clinically relevant given the reported association between unavailable cytogenetic results and poor prognosis in AML.^51^ WGS captured all clinically relevant SVs identified by SoC and detected additional cryptic rearrangements in four cases (4%), including one *NUP98*-, two *MECOM*- and one *KMT2A*-associated events. For clinically relevant CNAs, 47 of 61 events (77%) were concordant between WGS and SoC, while seven (11%) were detected only by WGS. The remaining CNA discrepancies mainly reflected low tumor content or small subclones (three SoC-only cases of trisomy 8, one monosomy 20), and differences in resolution and nomenclature between G-banding and WGS, particularly for 17p abnormalities. One complex t(3;3)(q21;q26) required manual reinterpretation of the detected SVs, highlighting that specific breakpoint regions may require targeted review. Overall, WGS provided high-resolution, integrated representation of SVs and CNAs compared with conventional cytogenetics, with breakpoint-level resolution and detection of submicroscopic alterations.

By integrating high-resolution CNA, CN-LOH, SV, and SNV data from WGS, the allelic state of *TP53* could be determined with greater precision than by SoC. This confirmed that most AMLs with *TP53* mutations in our cohort (12/13) involved both alleles, in line with recent genomic findings.^52^ This integrated assessment also reinforced the strong relationship between biallelic *TP53* disruption and genomic complexity that has been described using other methods.^45,47,52,53^ In fact, when measured by WGS, the number of detected chromosomal aberrations in bi*TP53* AML compared with other AMLs was significantly higher, highlighting bi*TP53* as a distinct subtype not only in MDS but potentially also in AML.^6^ Cytogenetically defined CK AML lacking a *TP53* mutation, however, displayed substantially fewer aberrations by WGS than bi*TP53* AML. This finding suggests that traditional CK definitions may group biologically heterogeneous entities and supports the view that *TP53* allelic state may represent a more fundamental determinant of genomic instability and clinical prognosis, whereas metaphase-based CK definitions, historically linked to adverse prognosis, may require re-evaluation.^47,48^

To provide a standardized, less labor-intensive, WGS-based assessment of genomic complexity, we developed the CK-probability score as an automated alternative to manual CK assessment. Although trained against cytogenetic CK, CK-prob classified all bi*TP53* AMLs as positive, including one case missed by SoC, while classifying fewer *TP53*-wildtype cases as positive compared with CK assessment by SoC. In our dataset, all CK *TP53*-wildtype cases harbored genetic abnormalities that take precedence over CK in the ICC classification hierarchy, showing a diminishing role of cytogenetically defined CK under the current classification frameworks. However, the proposed CK-prob will need to be evaluated in independent cohorts to prove its usability as a standardized automated diagnostic method for CK assessment.

Our data demonstrate that WGS has equal or better performance compared with SoC analyses, although some considerations for future clinical implementation remain. This retrospective study did not evaluate cost or turnaround time in routine diagnostics, which should be assessed prospectively. In addition, the WGS read depth employed in this study (90x), while higher than in previous studies^25^, has reduced sensitivity for variants with a VAF below 5%. However, ongoing advances in integrated single-platform workflows and sequencing technologies can reduce both turnaround times and costs of WGS,^54,55^ enabling higher sequencing depth and increased sensitivity for low-VAF variants.

In conclusion, these results support paired tumor–normal WGS as a single comprehensive assay for AML diagnostics, integrating disease classification, risk stratification, *TP53* allelic state evaluation, and automated assessment of CK.

## Acknowledgments

The authors thank Björn Hallström, Sailendra Pradhananga and Viktor Henmyr at the Section of Molecular Diagnostics, Skåne University Hospital, Lund, for support with the in-house bioinformatic pipeline. The authors also thank members of the SciLifeLab Clinical Genomics Platform and Genomic Medicine Sweden for their continued support in developing and implementing genomics-based methods in Swedish healthcare. Whole-genome sequencing was performed at the SNP&SEQ Technology Platform in Uppsala, which is part of the National Genomics Infrastructure Sweden and Science for Life Laboratory. The platform is supported by the Swedish Research Council and the Knut and Alice Wallenberg Foundation.

## Author contributions

**Petr Stanek:** Data curation; formal analysis; investigation; methodology; resources; software; visualization; writing – original draft; writing – review and editing. **Christina Orsmark-Pietras**: Data curation; conceptualization; investigation; resources; supervision; writing – review and editing. **Rasmus Henningsson:** Investigation; methodology; supervision; writing – review and editing. **Marianne Rissler:** Resources; writing – review and editing. **Niklas Landberg:** Data curation; writing – review and editing. **Helena Ågerstam:** Resources; writing – review and editing. **Anna Staffas:** Conceptualization; writing – review and editing. **Linda Fogelstrand:** Conceptualization; writing – review and editing. **Lars Palmqvist:** Conceptualization; writing – review and editing. **Gisela Barbany:** Conceptualization; writing – review and editing. **Mattis Knulst:** Data curation; Resources; writing – review and editing. **Gunnar Juliusson:** Resources; writing – review and editing. **Vladimir Lazarevic:** Resources; supervision; writing – review and editing. **Valtteri Wirta:** Conceptualization; funding acquisition; writing – review and editing. **Lucia Cavelier:** Conceptualization; funding acquisition; writing – review and editing. **Richard Rosenquist:** Conceptualization; funding acquisition; writing – review and editing. **Henrik Lilljebjörn:** Data curation; conceptualization; investigation; methodology; resources; software; supervision; writing – review and editing. **Thoas Fioretos:** Conceptualization; funding acquisition; investigation; project administration; resources; supervision; writing – review and editing.

## Data Availability Statement

Access to patient-level genomic data is subject to applicable ethical and data-protection regulations. Processed variant data supporting the findings of this study are provided in the Supplementary Tables S1-S6. Genomic data are available from the corresponding authors upon reasonable request and appropriate institutional approval.

## REFERENCES

1. Ley TJ, Mardis ER, Ding L, et al. DNA sequencing of a cytogenetically normal acute myeloid leukaemia genome. Nature. 2008;456(7218):66–72. doi:10.1038/nature07485

2. Ding L, Ley TJ, Larson DE, et al. Clonal evolution in relapsed acute myeloid leukaemia revealed by whole-genome sequencing. Nature. 2012;481(7382):506–510. doi:10.1038/nature10738

3. Papaemmanuil E, Gerstung M, Bullinger L, et al. Genomic Classification and Prognosis in Acute Myeloid Leukemia. New England Journal of Medicine. 2016;374(23):2209–2221. doi:10.1056/NEJMoa1516192

4. Döhner H, Wei AH, Appelbaum FR, et al. Diagnosis and management of AML in adults: 2022 recommendations from an international expert panel on behalf of the ELN. Blood. 2022;140(12):1345–1377. doi:10.1182/blood.2022016867

5. Arber DA, Orazi A, Hasserjian RP, et al. International Consensus Classification of Myeloid Neoplasms and Acute Leukemias: integrating morphologic, clinical, and genomic data. Blood. 2022;140(11):1200–1228. doi:10.1182/blood.2022015850

6. Khoury JD, Solary E, Abla O, et al. The 5th edition of the World Health Organization Classification of Haematolymphoid Tumours: Myeloid and Histiocytic/Dendritic Neoplasms. Leukemia. 2022;36(7):7. doi:10.1038/s41375-022-01613-1

7. DiNardo CD, Stein EM, Botton S de, et al. Durable Remissions with Ivosidenib in IDH1-Mutated Relapsed or Refractory AML. New England Journal of Medicine. 2018;378(25):2386–2398. doi:10.1056/NEJMoa1716984

8. Stein EM, DiNardo CD, Pollyea DA, et al. Enasidenib in mutant IDH2 relapsed or refractory acute myeloid leukemia. Blood. 2017;130(6):722–731. doi:10.1182/blood-2017-04-779405

9. Perl AE, Larson RA, Podoltsev NA, et al. Follow-up of patients with R/R FLT3-mutation–positive AML treated with gilteritinib in the phase 3 ADMIRAL trial. Blood. 2022;139(23):3366–3375. doi:10.1182/blood.2021011583

10. Issa GC, Aldoss I, DiPersio J, et al. The menin inhibitor revumenib in KMT2A-rearranged or NPM1-mutant leukaemia. Nature. 2023;615(7954):920–924. doi:10.1038/s41586-023-05812-3

11. Arber DA, Orazi A, Hasserjian R, et al. The 2016 revision to the World Health Organization classification of myeloid neoplasms and acute leukemia. Blood. 2016;127(20):2391-2405. doi:10.1182/blood-2016-03-643544

12. Döhner H, Estey E, Grimwade D, et al. Diagnosis and management of AML in adults: 2017 ELN recommendations from an international expert panel. Blood. 2017;129(4):424–447. doi:10.1182/blood-2016-08-733196

13. Duncavage EJ, Bagg A, Hasserjian RP, et al. Genomic profiling for clinical decision making in myeloid neoplasms and acute leukemia. Blood. 2022;140(21):2228–2247. doi:10.1182/blood.2022015853

14. Mrózek K, Heerema NA, Bloomfield CD. Cytogenetics in acute leukemia. Blood Reviews. 2004;18(2):115–136. doi:10.1016/S0268-960X(03)00040-7

15. Lee HR, Oh B, Hong DS, et al. Cytogenetic features of 5q deletion and 5q− syndrome in myelodysplastic syndrome in Korea; marker chromosomes proved to be chromosome 5 with interstitial deletion by fluorescence in situ hybridization. Cancer Genetics and Cytogenetics. 2010;203(2):193–202. doi:10.1016/j.cancergencyto.2010.08.007

16. Puente XS, Beà S, Valdés-Mas R, et al. Non-coding recurrent mutations in chronic lymphocytic leukaemia. Nature. 2015;526(7574):519–524. doi:10.1038/nature14666

17. Aaltonen LA, Abascal F, Abeshouse A, et al. Pan-cancer analysis of whole genomes. Nature. 2020;578(7793):82–93. doi:10.1038/s41586-020-1969-6

18. Alexandrov LB, Nik-Zainal S, Wedge DC, et al. Signatures of mutational processes in human cancer. Nature. 2013;500(7463):415–421. doi:10.1038/nature12477

19. Nik-Zainal S, Alexandrov LB, Wedge DC, et al. Mutational Processes Molding the Genomes of 21 Breast Cancers. Cell. 2012;149(5):979–993. doi:10.1016/j.cell.2012.04.024

20. Brady SW, Roberts KG, Gu Z, et al. The genomic landscape of pediatric acute lymphoblastic leukemia. Nat Genet. 2022;54(9):1376–1389. doi:10.1038/s41588-022-01159-z

21. Voss RK, Pastor Loyola VB, Cardenas MF, et al. Clinical experience of using integrated whole genome and transcriptome sequencing as a framework for pediatric and adolescent acute myeloid leukemia diagnosis and risk assessment. Leukemia. 2025;39(12):2946–2956. doi:10.1038/s41375-025-02774-5

22. Schmitz R, Wright GW, Huang DW, et al. Genetics and Pathogenesis of Diffuse Large B-Cell Lymphoma. New England Journal of Medicine. 2018;378(15):1396–1407. doi:10.1056/NEJMoa1801445

23. Wadensten E, Wessman S, Abel F, et al. Diagnostic Yield From a Nationwide Implementation of Precision Medicine for all Children With Cancer. JCO Precis Oncol. 2023;7:e2300039. doi:10.1200/PO.23.00039

24. Rezayee F, Eisfeldt J, Skaftason A, et al. Feasibility to use whole-genome sequencing as a sole diagnostic method to detect genomic aberrations in pediatric B-cell acute lymphoblastic leukemia. Front Oncol. 2023;13:1217712. doi:10.3389/fonc.2023.1217712

25. Duncavage EJ, Schroeder MC, O’Laughlin M, et al. Genome Sequencing as an Alternative to Cytogenetic Analysis in Myeloid Cancers. N Engl J Med. 2021;384(10):924–935. doi:10.1056/NEJMoa2024534

26. Huber S, Baer C, Hutter S, et al. AML classification in the year 2023: How to avoid a Babylonian confusion of languages. Leukemia. 2023;37(7):1413–1420. doi:10.1038/s41375-023-01909-w

27. Berglund E, Barbany G, Orsmark-Pietras C, et al. A Study Protocol for Validation and Implementation of Whole-Genome and -Transcriptome Sequencing as a Comprehensive Precision Diagnostic Test in Acute Leukemias. Front Med (Lausanne). 2022;9:842507. doi:10.3389/fmed.2022.842507

28. Fioretos T, Wirta V, Cavelier L, et al. Implementing precision medicine in a regionally organized healthcare system in Sweden. Nat Med. 2022;28(10):1980–1982. doi:10.1038/s41591-022-01963-4

29. Orsmark-Pietras C, Landberg N, Lorenz F, et al. Clinical and genomic characterization of patients diagnosed with the provisional entity acute myeloid leukemia with BCR-ABL1, a Swedish population-based study. Genes Chromosomes Cancer. 2021;60(6):426–433. doi:10.1002/gcc.22936

30. Orsmark-Pietras C, Lyander A, Ladenvall C, et al. Precision Diagnostics in Myeloid Malignancies: Development and Validation of a National Capture-Based Gene Panel. *Genes*, Chromosomes and Cancer. 2024;63(7):e23257. doi:10.1002/gcc.23257

31. Lilljebjörn H, Peña-Martínez P, Thorsson H, et al. The AML cellular state space unveils NPM1 immune evasion subtypes with distinct clinical outcomes. Nat Commun. 2025;16(1):10592. doi:10.1038/s41467-025-66546-6

32. Pradhananga S, Henmyr V, Hallström B. SMD-Bioinformatics-Lund/nextflow_tumwgs. Published online 2021. Accessed June 29, 2026. https://github.com/SMD-Bioinformatics-Lund/nextflow_tumwgs/tree/cfab2a16c097d41c5c55b59a6866d97837f88bf5

33. Garrison E, Marth G. Haplotype-based variant detection from short-read sequencing. arXiv. Preprint posted online July 20, 2012: arXiv:1207.3907. doi:10.48550/arXiv.1207.3907

34. Lai Z, Markovets A, Ahdesmaki M, et al. VarDict: a novel and versatile variant caller for next-generation sequencing in cancer research. Nucleic Acids Res. 2016;44(11):e108. doi:10.1093/nar/gkw227

35. Chen X, Schulz-Trieglaff O, Shaw R, et al. Manta: rapid detection of structural variants and indels for germline and cancer sequencing applications. Bioinformatics. 2016;32(8):1220–1222. doi:10.1093/bioinformatics/btv710

36. McKenna A, Hanna M, Banks E, et al. The Genome Analysis Toolkit: a MapReduce framework for analyzing next-generation DNA sequencing data. Genome Res. 2010;20(9):1297–1303. doi:10.1101/gr.107524.110

37. McLaren W, Gil L, Hunt SE, et al. The Ensembl Variant Effect Predictor. Genome Biol. 2016;17(1):122. doi:10.1186/s13059-016-0974-4

38. Danecek P, Bonfield JK, Liddle J, et al. Twelve years of SAMtools and BCFtools. Gigascience. 2021;10(2):giab008. doi:10.1093/gigascience/giab008

39. Miller CA, Walker JR, Jensen TL, et al. Failure to Detect Mutations in *U2AF1* due to Changes in the GRCh38 Reference Sequence. The Journal of Molecular Diagnostics. 2022;24(3):219–223. doi:10.1016/j.jmoldx.2021.10.013

40. Pedregosa F, Varoquaux G, Gramfort A, et al. Scikit-learn: Machine Learning in Python. Journal of Machine Learning Research. 2011;12(85):2825–2830.

41. Virtanen P, Gommers R, Oliphant TE, et al. SciPy 1.0: fundamental algorithms for scientific computing in Python. Nat Methods. 2020;17(3):261–272. doi:10.1038/s41592-019-0686-2

42. Yannakou CK, Jones K, McBean M, et al. ASXL1 c.1934dup;p.Gly646Trpfs*12—a true somatic alteration requiring a new approach. Blood Cancer Journal. 2017;7(12):656. doi:10.1038/s41408-017-0025-8

43. Xie DH, Chen WM, Hao Y, et al. The Characteristics and Outcomes of Adult Acute Myeloid Leukemia Patients With KMT2A-Partial Tandem Duplication. International Journal of Laboratory Hematology. 2025;47(6):1119–1127. doi:10.1111/ijlh.14532

44. Eriksson A, Engvall M, Mathot L, et al. Somatic Exonic Deletions in RUNX1 Constitutes a Novel Recurrent Genomic Abnormality in Acute Myeloid Leukemia. Clin Cancer Res. 2023;29(15):2826–2834. doi:10.1158/1078-0432.CCR-23-0122

45. Rücker FG, Schlenk RF, Bullinger L, et al. TP53 alterations in acute myeloid leukemia with complex karyotype correlate with specific copy number alterations, monosomal karyotype, and dismal outcome. Blood. 2012;119(9):2114–2121. doi:10.1182/blood-2011-08-375758

46. Seifert H, Mohr B, Thiede C, et al. The prognostic impact of 17p (p53) deletion in 2272 adults with acute myeloid leukemia. Leukemia. 2009;23(4):656–663. doi:10.1038/leu.2008.375

47. Stengel A, Meggendorfer M, Walter W, et al. Interplay of TP53 allelic state, blast count, and complex karyotype on survival of patients with AML and MDS. Blood Adv. 2023;7(18):5540–5548. doi:10.1182/bloodadvances.2023010312

48. Weinberg OK, Siddon A, Madanat YF, et al. TP53 mutation defines a unique subgroup within complex karyotype de novo and therapy-related MDS/AML. Blood Adv. 2022;6(9):2847–2853. doi:10.1182/bloodadvances.2021006239

49. Hart SA, Lee LA, Seegmiller AC, Mason EF. Diagnosis of *TP53*-mutated myeloid disease by the ICC and WHO fifth edition classifications. Blood Advances. 2025;9(3):445–454. doi:10.1182/bloodadvances.2024014140

50. Cancer Genome Atlas Research Network, Ley TJ, Miller C, et al. Genomic and epigenomic landscapes of adult de novo acute myeloid leukemia. N Engl J Med. 2013;368(22):2059–2074. doi:10.1056/NEJMoa1301689

51. Lazarevic V, Hörstedt AS, Johansson B, et al. Failure matters: unsuccessful cytogenetics and unperformed cytogenetics are associated with a poor prognosis in a population-based series of acute myeloid leukaemia. European Journal of Haematology. 2015;94(5):419–423. doi:10.1111/ejh.12446

52. Abel HJ, Oetjen KA, Miller CA, et al. Genomic landscape of TP53-mutated myeloid malignancies. Blood Adv. 2023;7(16):4586–4598. doi:10.1182/bloodadvances.2023010156

53. Kim K, Maiti A, Loghavi S, et al. Outcomes of TP53-mutant acute myeloid leukemia with decitabine and venetoclax. Cancer. 2021;127(20):3772–3781. doi:10.1002/cncr.33689

54. Abel HJ, Mahgoub M, Davarapalli N, et al. Evaluation of Long-Read Genome Sequencing for Genomic Profiling of Myeloid Cancers. J Mol Diagn. 2025;27(12):1242–1254. doi:10.1016/j.jmoldx.2025.09.001

55. Vedi A, Trotman J, Dias JM, et al. Clinical Impact of Ultra-Fast Whole Genome Sequencing in Paediatric Haematology-Oncology Practice. medRxiv. Preprint posted online September 2, 2025:2025.08.29.25334714. doi:10.1101/2025.08.29.25334714

