## Supplemental Figures for "Whole-genome sequencing enables improved AML diagnostics and refines assessment of *TP53* allelic state and complex karyotype"

**Figure S1**

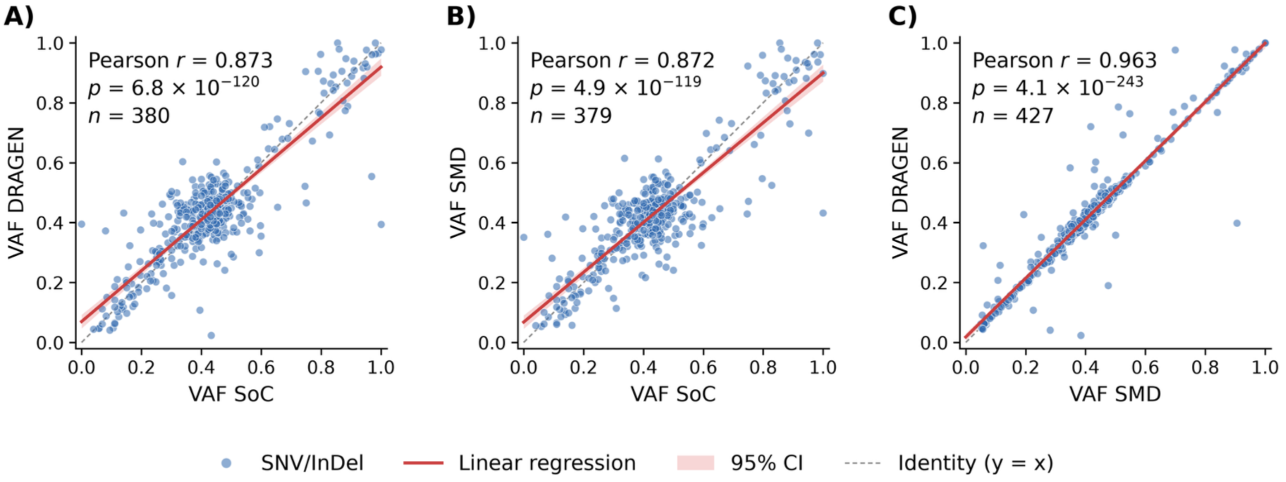


**Figure S1. Concordance of variant allele frequencies between SoC and WGS pipelines.**

Red lines indicate fitted linear regressions; shaded bands indicate 95% confidence interval (CI). The Pearson correlation coefficient associated P value, and the total number of paired measurements (n) is shown in each panel.

Abbreviations: InDel, insertion/deletion; SMD, Section of Molecular Diagnostics (in-house WGS analysis pipeline); SNV, single-nucleotide variant; SoC, standard-of-care; VAF, variant allele frequency; WGS, whole-genome sequencing.

### Figure S2

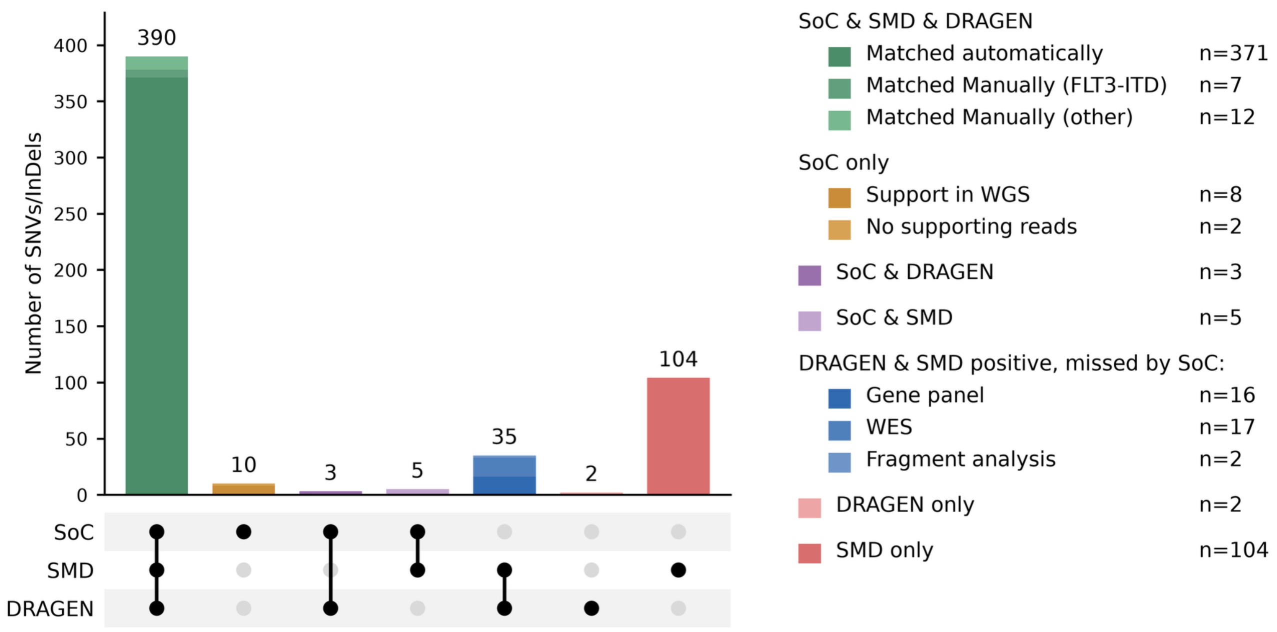


**Figure S2. Overlap of SNV and InDel calls between SoC and the two WGS pipelines.**

Upset plot showing variants detected by SoC, WGS-SMD, and WGS-DRAGEN. Connected black dots identify the methods contributing to each intersection; bars indicate the corresponding variant counts. Variants shared by all three methods are subdivided into automatic matches and manual matches involving *FLT3* internal tandem duplications or other variants, as indicated by color shade. SoC-only calls are colored according to the presence or absence of supporting WGS reads. WGS-only calls are colored based on which corresponding SoC assay was performed (i.e. which SoC assay failed to detect the variant: gene panel, whole-exome sequencing, or fragment analysis). Calls unique to one WGS pipeline are presented separately. All variants missed solely by WGS-DRAGEN represent SNVs in high-homology region of *U2AF1*.

Abbreviations: DRAGEN, Dynamic Read Analysis for GENomics (commercial WGS analysis pipeline); InDel, insertion/deletion; ITD, internal tandem duplication; SMD, Section of Molecular Diagnostics (in-house WGS analysis pipeline); SNV, single-nucleotide variant; SoC, standard of care; WES, whole-exome sequencing; WGS, whole-genome sequencing.

### Figure S3

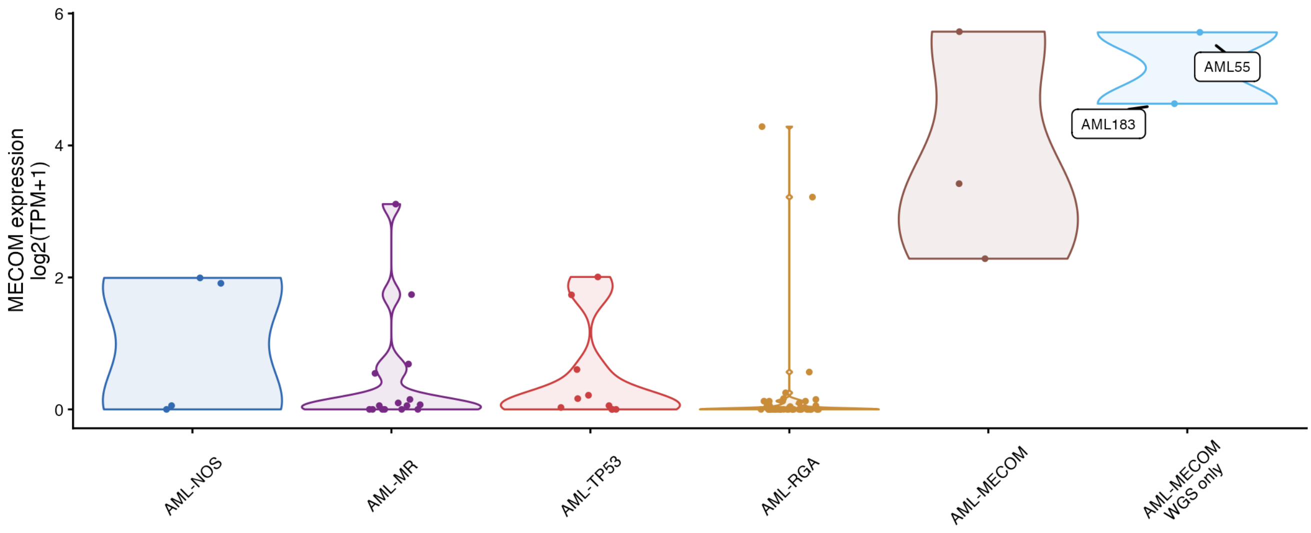


**Figure S3. *MECOM* expression in 83 AML samples with available RNA-sequencing data.**

Violin plots show the distribution of *MECOM* expression, with points representing individual samples. Samples are grouped by major ICC categories, with *MECOM*-rearranged cases shown separately according to detection by both SoC and WGS (AML-*MECOM*: AML44, AML154, and AML172) or by WGS only (AML-*MECOM* WGS-only: AML55 and AML183). RNA-sequencing data was not available for AML343, also harboring a *MECOM* rearrangement.

Abbreviations: AML, acute myeloid leukemia; ICC, International Consensus Classification; MR, myelodysplasia-related; NOS, not otherwise specified; RGA, recurrent genetic abnormality; SoC, standard of care; TPM, transcripts per million; WGS, whole-genome sequencing.

**Figure S4**

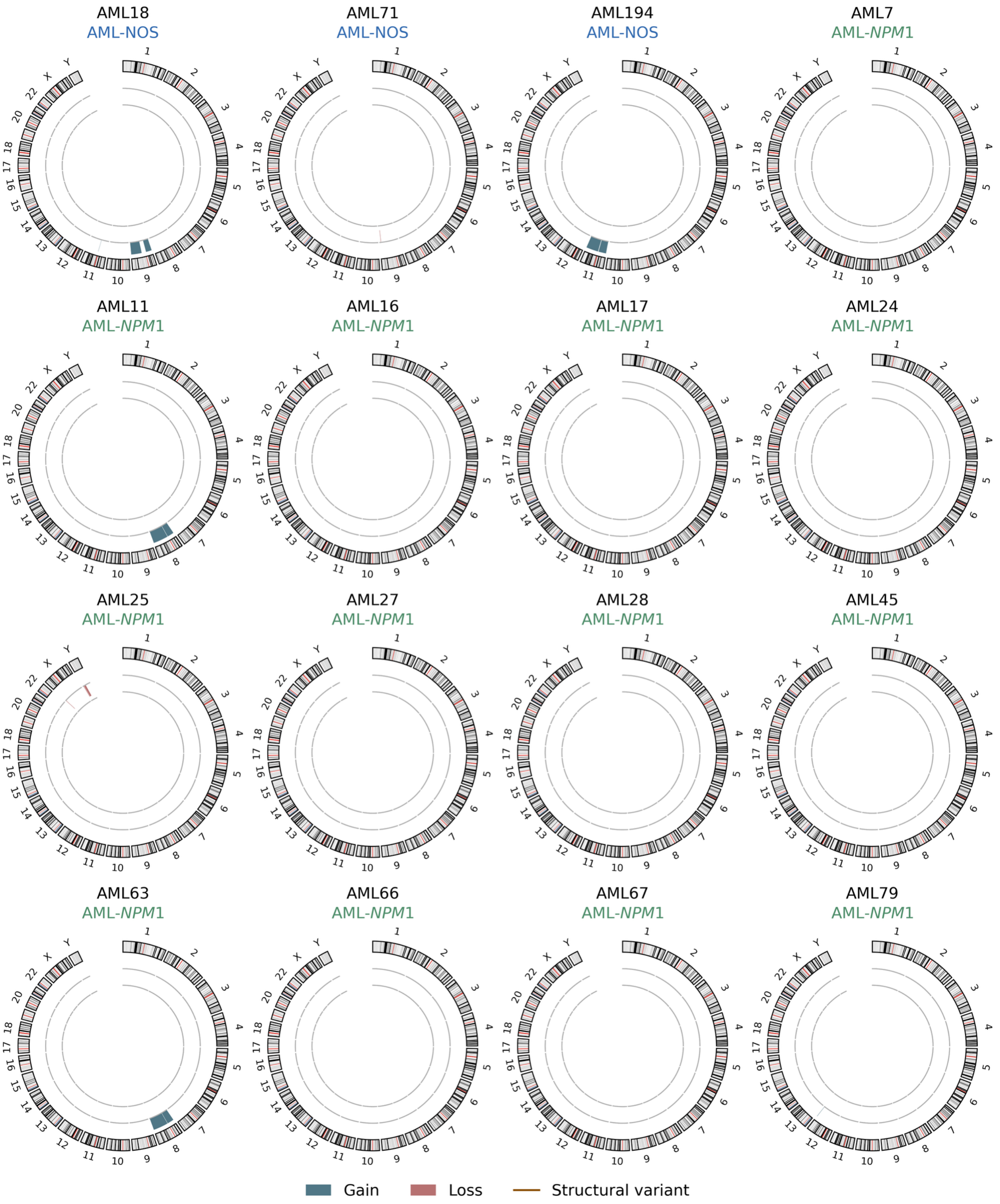


**Figure S4. Genome-wide structural and copy-number alterations across 16 AML-NOS and AML-*NPM1* samples.**

Circos plots showing WGS-detected alterations in three AML-NOS (AML not otherwise specified; AML18, AML71 and AML194) samples and thirteen AML-*NPM1* (AML with mutated *NPM1*; AML7 to AML79) samples. Subgroups reflect supplementary table S10.

### Figure S5

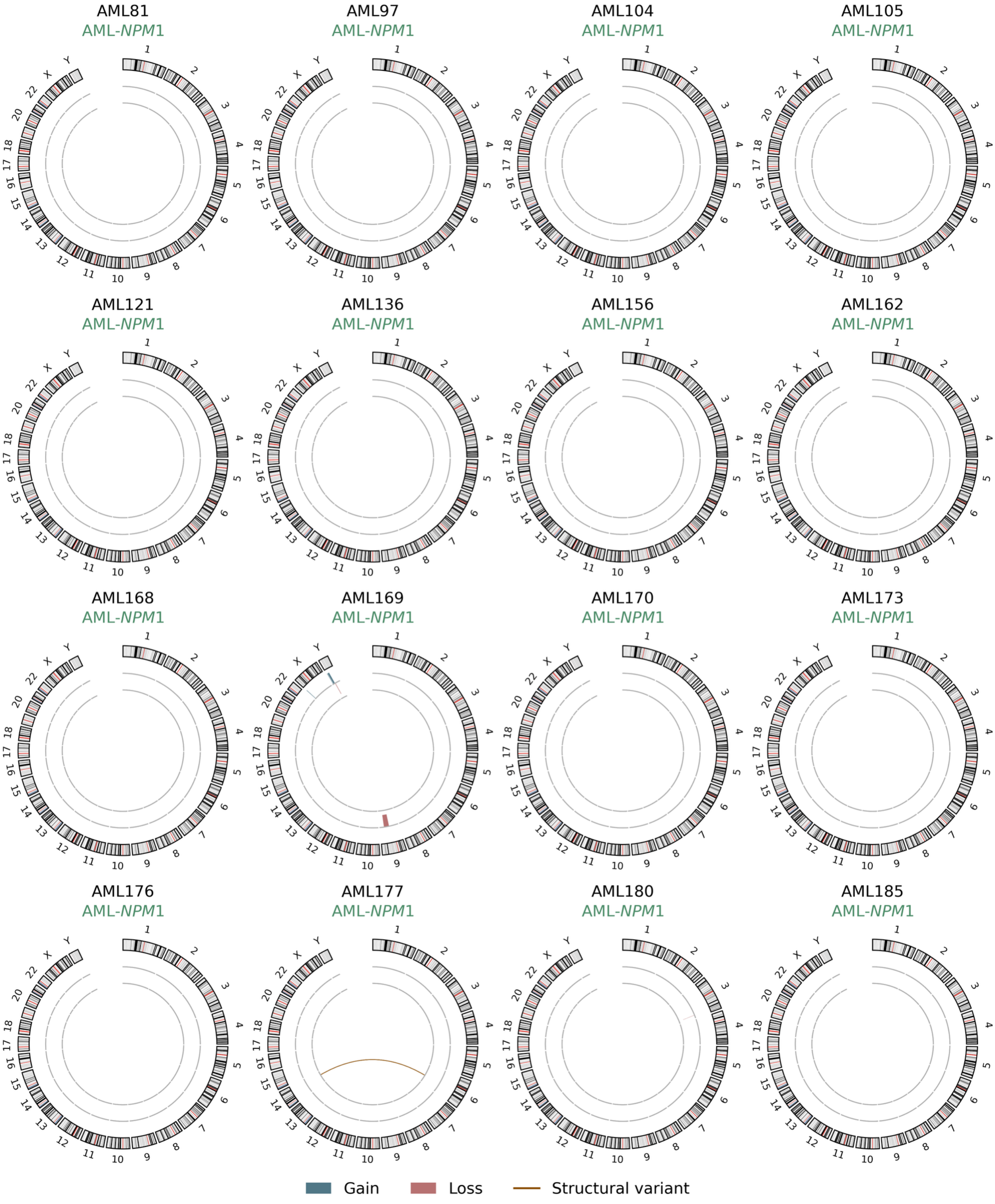


**Figure S5. Genome-wide structural and copy-number alterations across 16 AML-*NPM1* samples.**

Circos plots showing WGS-detected alterations in 16 AML-*NPM1* (AML with mutated *NPM1*; AML81 to AML185) samples. Subgroups reflect supplementary table S10.

**Figure S6**

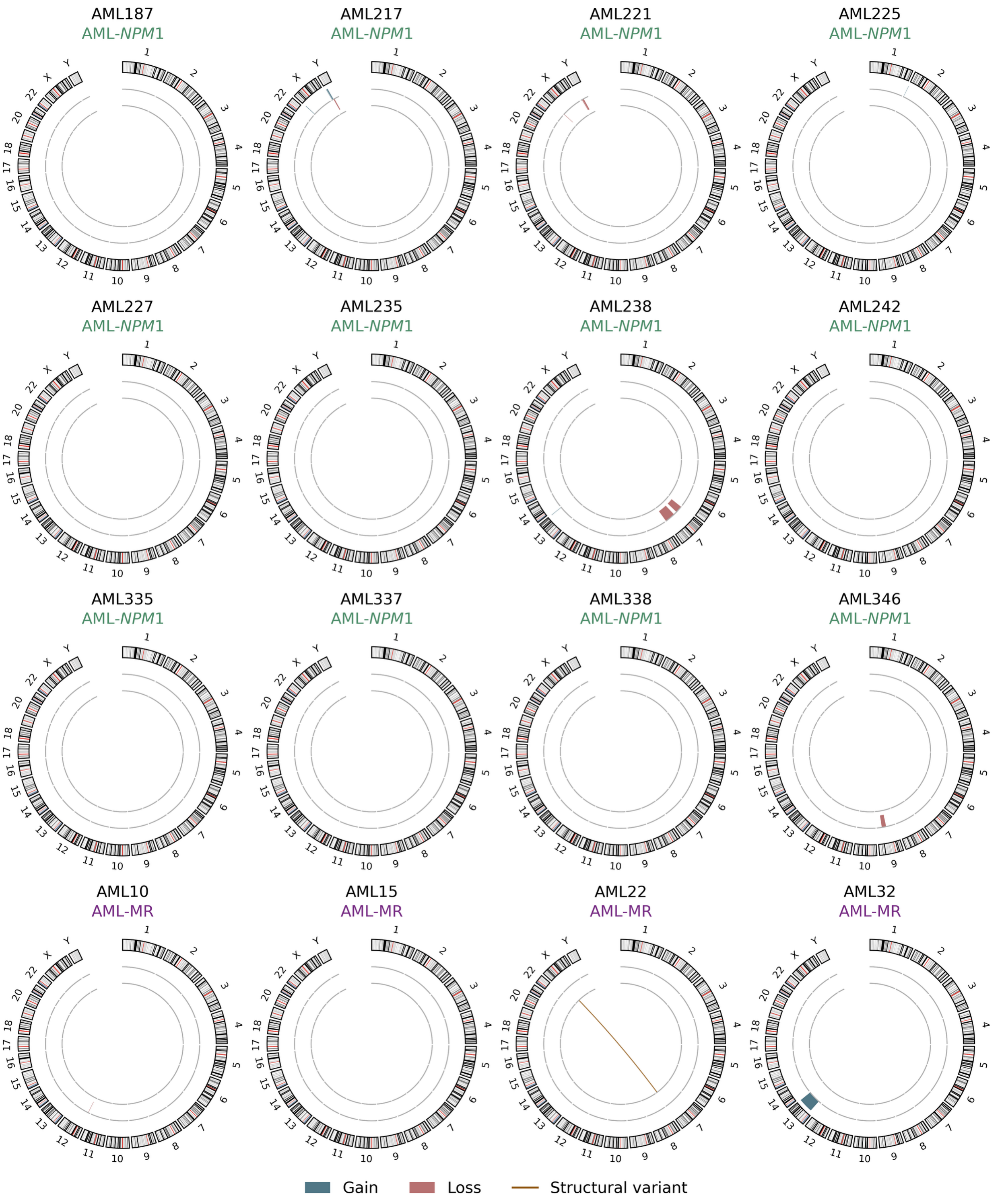


**Figure S6. Genome-wide structural and copy-number alterations across 16 AML-*NPM1* and AML-MR samples.**

Circos plots showing WGS-detected alterations in four AML-MR (AML with myelodysplasia-related gene mutations or cytogenetic abnormalities; AML10 to AML32) samples and twelve AML-*NPM1*

(AML with mutated *NPM1*; AML187 to AML346) samples. Subgroups reflect supplementary table S10.

**Figure S7**

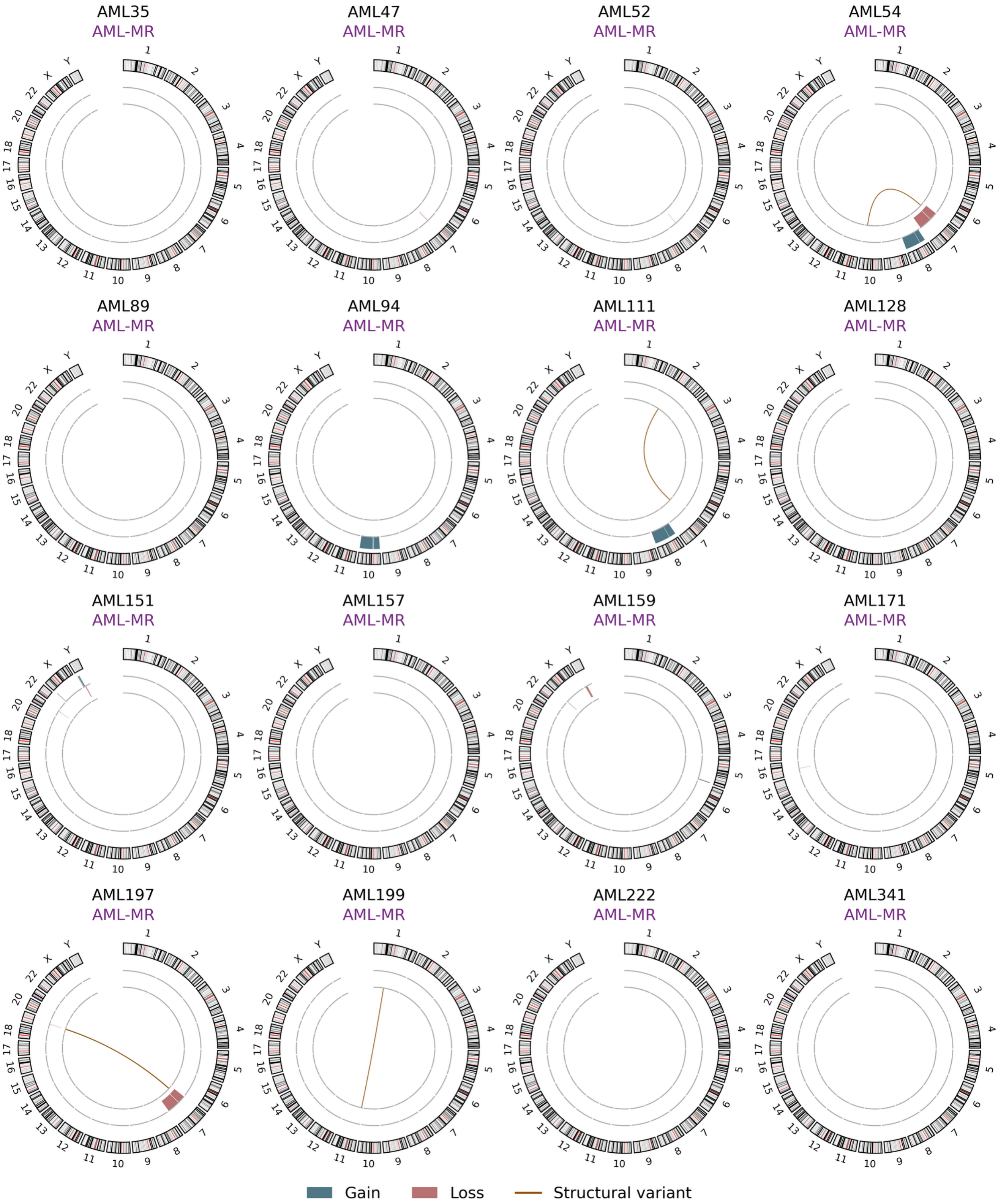


**Figure S7. Genome-wide structural and copy-number alterations across 16 AML-MR samples.**

Circos plots showing WGS-detected alterations in 16 AML-MR (AML with myelodysplasia-related gene mutations or cytogenetic abnormalities; AML35 to AML341) samples. Subgroups reflect supplementary table S10.

### Figure S8

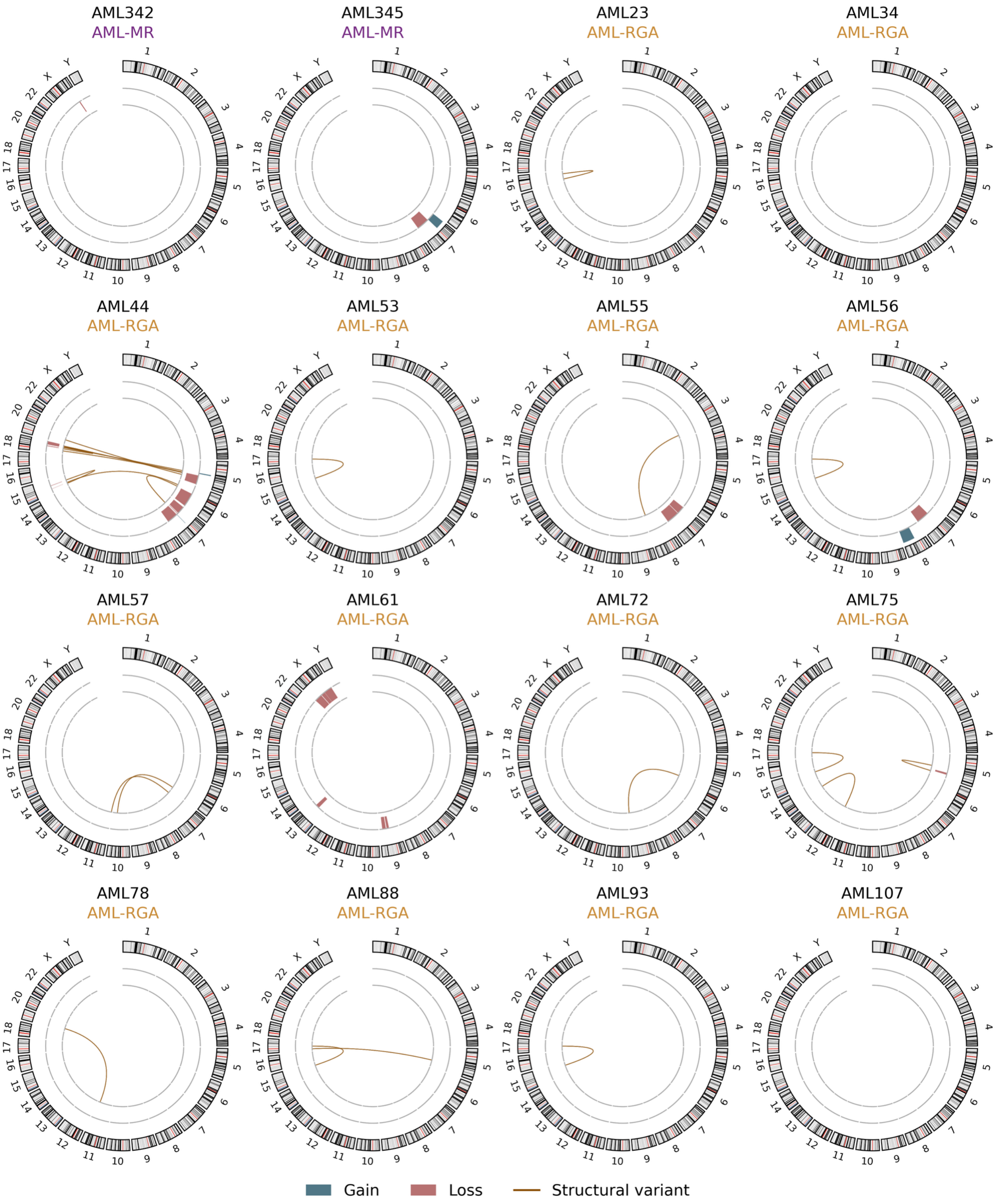


**Figure S8. Genome-wide structural and copy-number alterations across 16 AML-MR and AML-RGA samples.**

Circos plots showing WGS-detected alterations in twelve AML-RGA (AML with other recurrent genetic abnormalities; AML23 to AML107) and two AML-MR (AML with myelodysplasia-related gene mutations or cytogenetic abnormalities; AML342 and 345) samples. Subgroups reflect supplementary table S10.

**Figure S9**

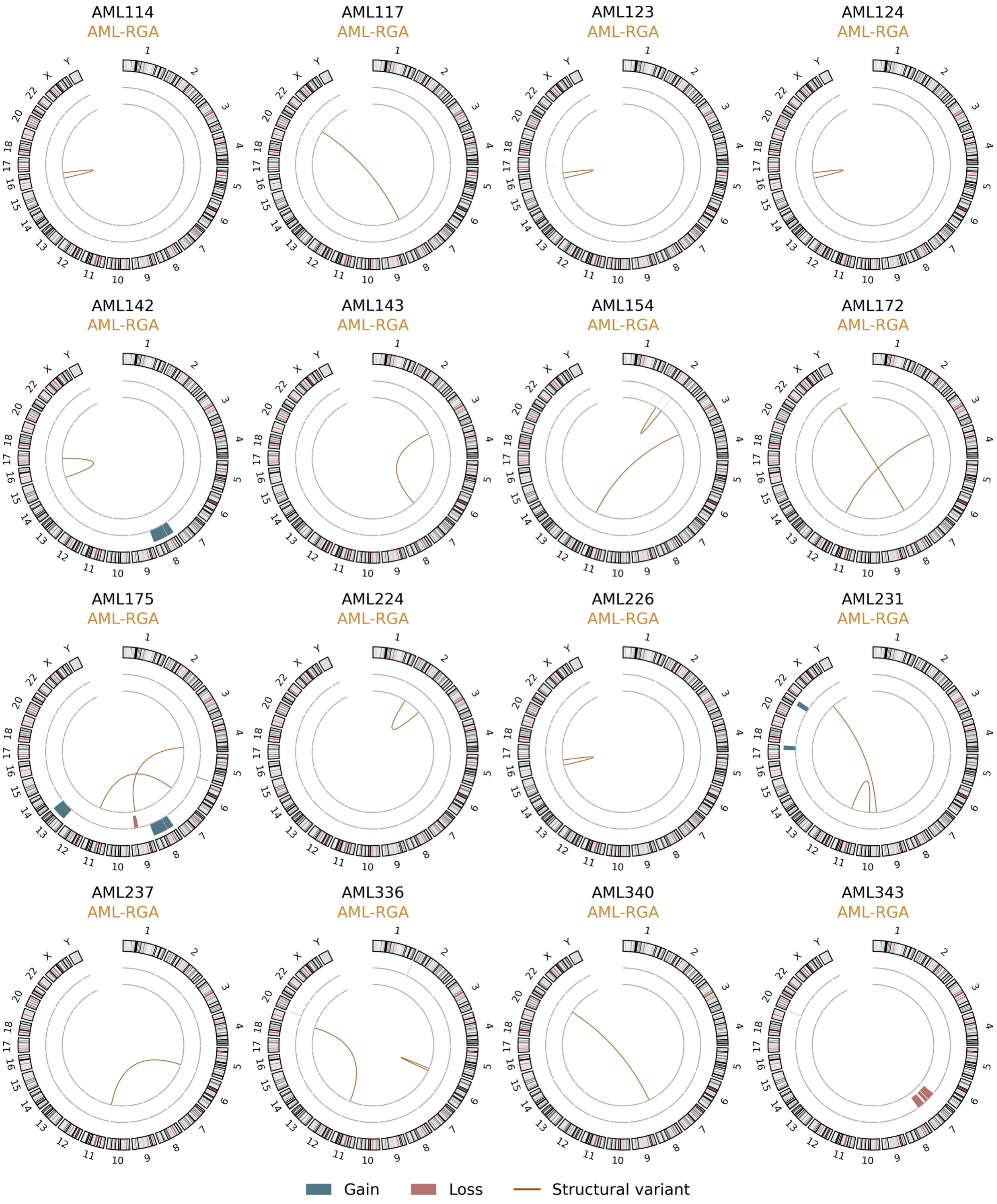


**Figure S9. Genome-wide structural and copy-number alterations across 16 AML-RGA samples.**

Circos plots showing WGS-detected alterations in 16 AML-RGA (AML with other recurrent genetic abnormalities; AML114 to AML343) samples. Subgroups reflect supplementary table S10.

**Figure S10**

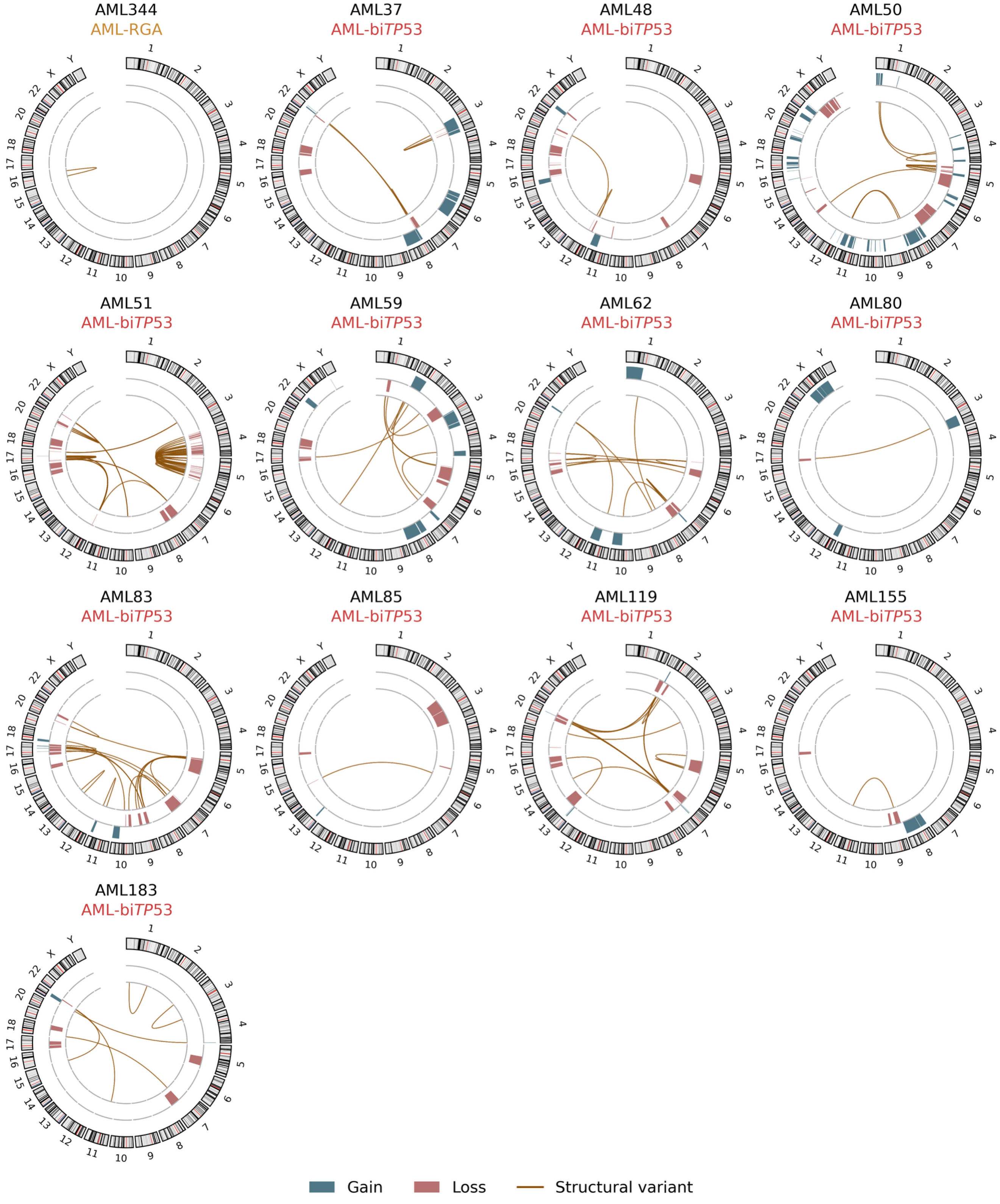


**Figure S10. Genome-wide structural and copy-number alterations across 13 AML-RGA and AML-bi*TP53* samples.**

Circos plots showing WGS-detected alterations in one AML-RGA (AML with other recurrent genetic abnormalities; AML344) and twelve AML-biTP53 (AML with biallelic *TP53* alterations; AML37 to AML183) samples. Subgroups reflect supplementary table S10. AML-bi*TP53* cases are grouped together irrespective of co-occurring class-defining abnormalities.

**Figure S11**

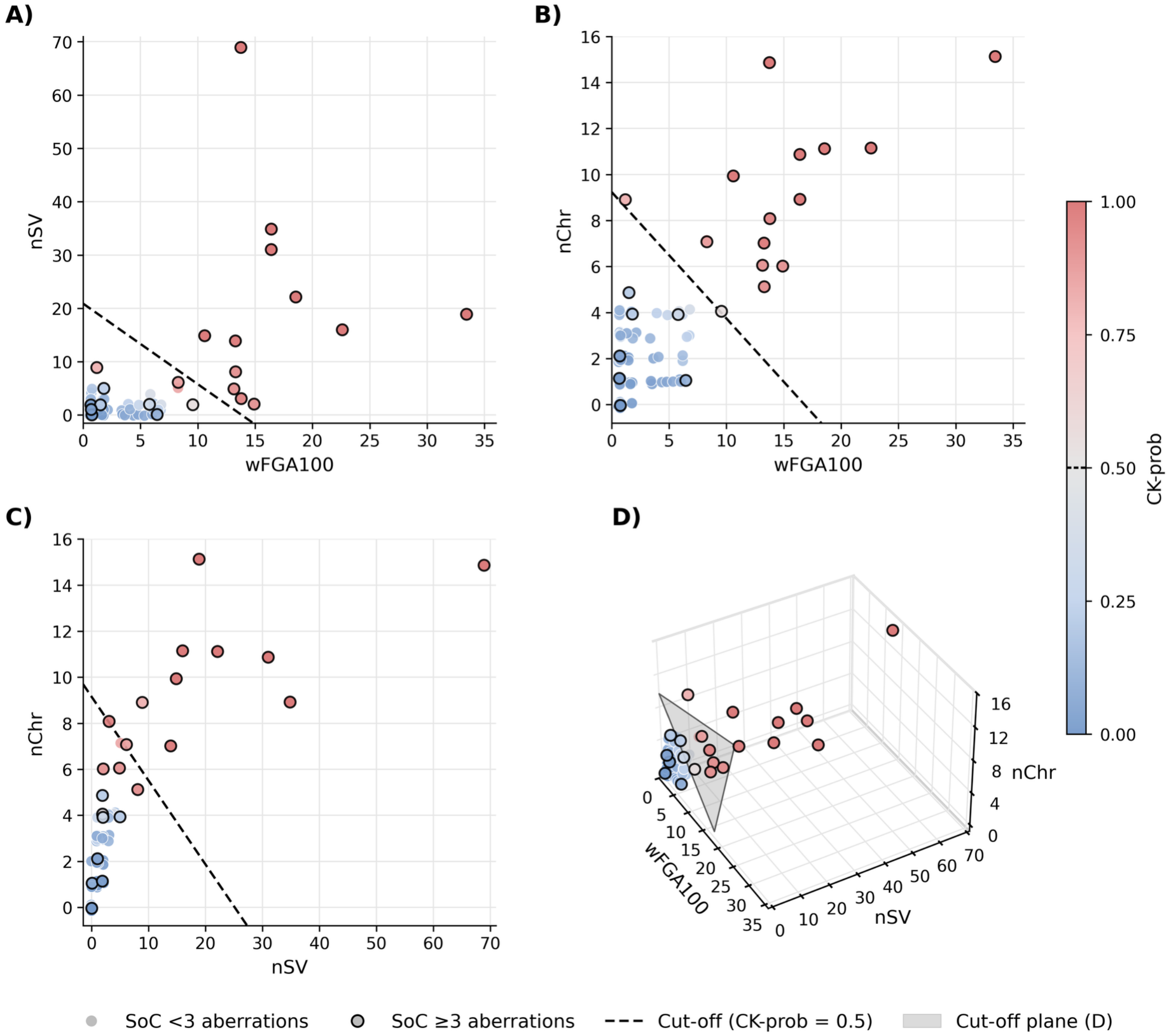


**Figure S11. WGS-derived predictors and decision boundaries of the CK-prob model.**

Each point represents a patient sample; color indicates CK-prob, and black outlines identify samples with ≥3 aberrations by SoC cytogenetics. (A) nSV versus wFGA, with a dashed lines indicating the boundary for CK-prob = 0.5 (boundary calculated assuming nChr=2, representing the median value over all samples). (B) nChr versus wFGA100, with a dashed lines indicating the boundary for CK-prob = 0.5 (boundary calculated assuming nSV=1, representing the median value over all samples). (C) nChr versus nSV, with a dashed lines indicating the boundary for CK-prob = 0.5 (boundary calculated assuming wFGA100=0.83, representing the median value over all samples). (D) Three-dimensional representation of the predictors; the gray plane indicates CK-prob = 0.5. Point colors reflect all three predictors and therefore need not match the side of a conditional boundary in panels A–C.

Abbreviations: CK, complex karyotype; CK-prob, complex karyotype probability score; CNA, copy-number alteration; nChr, number of chromosomes affected by a CNA spanning ≥5% of chromosome length or a high-confidence SV; nSV, number of high-confidence structural variants; SoC, standard of care; SV, structural variant; wFGA100, CN-weighted fraction of the genome affected by CNAs multiplied by 100 (wFGA100 in the main text); WGS, whole-genome sequencing.
